# Rare variant associations of chronic pain conditions in the UK Biobank Whole Exome Sequencing

**DOI:** 10.1101/2025.10.25.25338777

**Authors:** Aidan P Nickerson, Julian Mutz, Samuel Handelman, Ryan Smith, Gopuraja Dharmalingam, David Collier, Cathryn Lewis, James P Dunham, Anthony E Pickering, Achim Kless

**Author notes:** Correspondence to: Achim Kless, Full address: Discovery Neuroscience Research, Neuroscience Genetics, Eli Lilly and Company Ltd, 8 Arlington Square West, Downshire Way, Bracknell, RG12 1PU, UK.

## Abstract

**Introduction:** In this population-based genetic study of chronic pain, we performed association analysis, including rare variants analysis, across a broad spectrum of conditions. While it is well-established that genetic factors play a significant role in the susceptibility to chronic pain, genome-wide association studies (GWAS) conducted to date have only uncovered a small portion of the genetic variants that likely contribute to its heritability. Our study identified shared genetic contributors to pain conditions at variant and gene levels, on a genome-wide basis.

**Methods:** We performed genome wide association analysis of chronic pain using the whole exome sequencing data in the UK Biobank across chronic pain conditions using optimized control cohorts. 355 chronic pain endpoints were derived from ICD diagnoses and a pain-specific questionnaire. For that purpose, we recalibrated principal components to account for population stratification and designed control cohorts to minimize bias and variability in the identified associations. Additionally, we analysed the shared genetic heritability of these different pain conditions using latent causal variable analysis on genotypic array data. Finally, we compared genes from our genome-wide significant associations with prior reported pain relevant genes.

**Results:** We found novel associations for mutations in all investigated pain conditions, with a particular emphasis on rare variants in the full UK Biobank cohort, which opens new opportunities for therapeutic approaches in the age of genetic medicines. Consequently, novel mechanisms of enigmatic diseases like complex regional pain syndrome and Fibromyalgia can be proposed. Many of these associations revealed the relevance of auto-immune processes and vascular pathophysiology. For example, in complex regional pain syndrome we found several novel genome wide significant associations. Examples include a causal rare missense variant in sialic acetyl esterase (SIAE R479C, p-value 3.34E-08, odds ratio 7.1, MAF 1e-04) and a protective rare missense variant in Mammary Analogous Zinc Finger 2 (MAML2, p-value 6.57E-06, odds ratio 0.62, MAF 0.015, I480M, rs61749251), which acts as a transcriptional cofactor for Notch proteins.

We conducted epistasis analyses on top-ranking variants to assess mutation penetrance and identified a causal mutation in *JCAD* (V366E; β = 3.4, χ² = 20.4, OR = 29.9, p = 6.39×10⁻⁶), a gene previously implicated in coronary artery disease. Given JCAD’s role in endothelial cell junctions and vascular integrity, this mutation may also influence neurovascular contributions to pain signalling, particularly in chronic inflammatory states.

Importantly, we discovered a rare protective variant in *PLA2G7* (S388P; β = –2.1, OR = 0.12, χ² = 12.9, p = 3.16×10⁻⁴), encoding lipoprotein-associated phospholipase A2, which is involved in phospholipid catabolism during inflammation and oxidative stress. This variant appears to limit oxylipin accumulation in the vascular wall—a mechanism that may reduce neuroinflammation and peripheral sensitization, offering a protective effect against pain pathophysiology.

The use of several control groups of different sizes allowed us to analyse the variability of results depending on this control cohort composition to characterize the sensitivity of genetic associations. This revealed several novel genome wide significant associations that were formerly of lower significance in osteoarthritis. Within our dataset of associations, we found frequently occurring variants that were associated with multiple chronic pain phenotypes suggesting common mechanisms across different pain conditions. Finally, we highlight the identification of novel mutations in known pain genes especially in sodium, potassium and calcium channels.

**Conclusions:** This study provides valuable insights into the genetic architecture of pain conditions by leveraging rare variant analysis and using optimized controls in a large-scale population cohort. Our findings implicate novel genetic associations and their role in pain susceptibility, paving the way for targeted research in the understanding of chronic pain conditions.

## Introduction

Chronic pain is a complex disease with varying aetiologies, clinical manifestations, and impacts on the individual’s life which summate to enormous implications for societies worldwide. While there are major environmental factors that predispose to or cause chronic pain, such as injury, there is also a strong inherited genetic component that influences its manifestations[1]. Targeted genetic studies of pain have identified rare monogenic disease-causing variants within families that have strong pain phenotypes such as complete insensitivity [2, 3] or patterns of extreme pain like Erythromelalgia [4]. These studies perform deep sequencing within families on a small number of genes within known nociceptive pathways with subsequent functional characterization using in-vitro, ex-vivo systems or even non-interventional clinical trials [2, 5]. These methods depend on the hypothesis that chronic pain is caused by a small number of pathogenic variants in known genes directly impacting signal transduction mechanisms typically influencing nociceptor excitability or the development of the sensory nervous system. While these studies have furthered the field of pain research, the general translation into subsequently derived therapeutics has yet to be shown.

The role of common genetic variation of complex traits is becoming more well-characterized as consortia using genome wide association studies (GWAS) have expanded to include hundreds of thousands of participants including in UK Biobank[6]. Such population-based cohorts also allow the investigation of rare variants based on the presence of identified carriers. For most of the chronic pain endpoints, GWAS have been performed at varying levels of DNA sequencing, control groups, and disease definitions. Specific conditions like fibromyalgia (FMS) [7], migraine [8, 9], osteoarthritis (OA)[10], rheumatoid arthritis (RA)[11] and complex regional pain syndrome (CRPS) [12] revealed numerous potentially relevant mutations and disease mechanisms (for systematic literature review see [13]). Recent efforts have compiled databases with genetic evidence across different genetic study designs and pain conditions into single resources [14]. Additionally, consortia and large institutes are using advanced data analytics to identify genetic associations across multiple diseases, such as OpenTargets[15] and OMIM[16]. These resources highlight the disparate signals across the approaches, with rather few genes involved in the nociceptive pathways identified by these GWAS, and the hits are often more closely linked to known pain co-morbid conditions like psychiatric diseases and obesity [17].

One further limitation is that many of these studies are using genotype arrays which can detect a limited number of variants at predefined positions. Such genotyping arrays typically use common variants across the entire genome, at the expense of rare variants like non-synonymous variants that can directly impact the function of the encoded protein. It is well established that those rare variants are more likely to have a large, clinically relevant effect on phenotypes and clinical presentation[18]. However, the rarity of these variants requires a very large sample size to have adequate statistical power to detect effects in fewer carriers [19]. Advances in genetic sequencing have enabled large scale studies of coding regions using whole exome sequencing. The UK Biobank recently released the whole exome sequencing of half a million individuals [20] including deep phenotyping based on electronic health records, imaging and blood based biomarkers as well as novel questionnaires like the pain questionnaire [21].

The complexity of chronic pain is hard to capture in most case-control based genetic studies since either a single pain condition was investigated in isolation, or cases have been defined by a single instrument. Most importantly, control group definitions are not rigorously defined. One example [22] is based on using the question: “In the last 3 months what area has been the most painful”, those that report knee pain are assigned in the “case” group, and those that report “None” are assigned to the control. However the high prevalence of chronic pain, estimated 35-51% in the UK[23], as well as a high incidence of comorbidities like anxiety, depression, obesity and other chronic pain conditions [24–26] may result in control groups also containing individuals with genetic variants that are influencing risk and progression of chronic pain. It has been formally demonstrated that incorporating incorrectly classified controls in genetic studies, particularly when there are significant misclassification rates, has a substantial impact [27].This misclassification problem is particularly relevant for rare variants where small numbers may have large impact upon the assigned significance [28].

In this large-scale genetic study of pain, we perform genetic association analysis in the whole exome sequencing dataset across a broad spectrum of chronic pain conditions using an optimized ‘pain free’ control cohort to account for the high prevalence and diversity of the clinical presentation of chronic pain in the general population. Further, we leveraged these associations to identify the shared genetic contributors and modifiers at the variant and gene levels on a genome-wide basis.

## Methods

### Pain Cohort definitions

We derived cohorts of participants with chronic pain conditions from pain relevant ICD9 and ICD10 codes (fields 41271 and 41270 respectively), initial interview responses (field 20002 – [INT] in phenotype name), experience of pain questionnaire (field category 154 - [PQ] in phenotype name), procedures for chronic pain (field 41272 – [OP] in phenotype name) and medication data (field 20003).

Given the hierarchical layout of ICD10 codes, cohorts were defined at both the most detailed level possible and at broader grouped nodes in cases where *N*<100 in subgroups e.g. G60 (Hereditary and idiopathic neuropathy) instead of G60.x. Only cohorts where N>100 were considered for genetic association analysis as per standard practice, allowing for larger effects to be observed [29]. Further we diversified in case of higher numbers like a cohort of “Vertebrogenic low back pain” was defined using the ICD10 code M54.51 as well as the broader group of “lower back pain” which includes M54.51 and other subgroups “Low back pain unspecified” (M54.50), and “Other low back pain” (M54.49). Additional questionnaire summary metrics were calculated from the experience of pain questionnaire. A “Douleur Neuropathique en Quatre” (DN4)[30] score was calculated using a sum of all “yes” responses in fields 120046-120051, considering a neuropathic score as greater than or equal to 3. We also defined cohorts by combinations of endpoints such as a diagnostic derived painful diabetic polyneuropathy with a diagnosis of diabetes (E11) together with a DN4 score greater than 3 from the pain questionnaire.

In total 329 pain derived phenotypes were analysed; full details of the cohorts and summary statistics of all genetic associations are available in the supplementary material. In this publication, we focus on a subset of phenotypes with known links to chronic pain. This includes self-reported phenotypes determined by initial interview (field 20002) : Unspecified pain for greater than 3 months (Chronic pain unspecified), Abdominal pain, Back pain, Joint pain, Carpal tunnel syndrome (CTS), Knee pain and Trigeminal neuralgia (TGN); Self-reported determined by pain questionnaire (UK biobank category 154): Rheumatoid arthritis, Cancer pain, Pelvic pain, complex regional pain syndrome (CRPS), post herpetic neuralgia (PHN) and fibromyalgia (FMS); hospital administration recorded ICD codes: Seronegative Rheumatoid arthritis (RA seronegative); Seropositive Rheumatoid arthritis (RA seropositive), Migraine, Low back pain, Diabetic polyneuropathy (DPN) and Idiopathic neuropathy; and by hospital procedures coded in OPCS4: Nerve destruction surgery and Joint replacement of hip or knee. Demographic features of these cohorts are shown in Table 1.

**Table 1:**
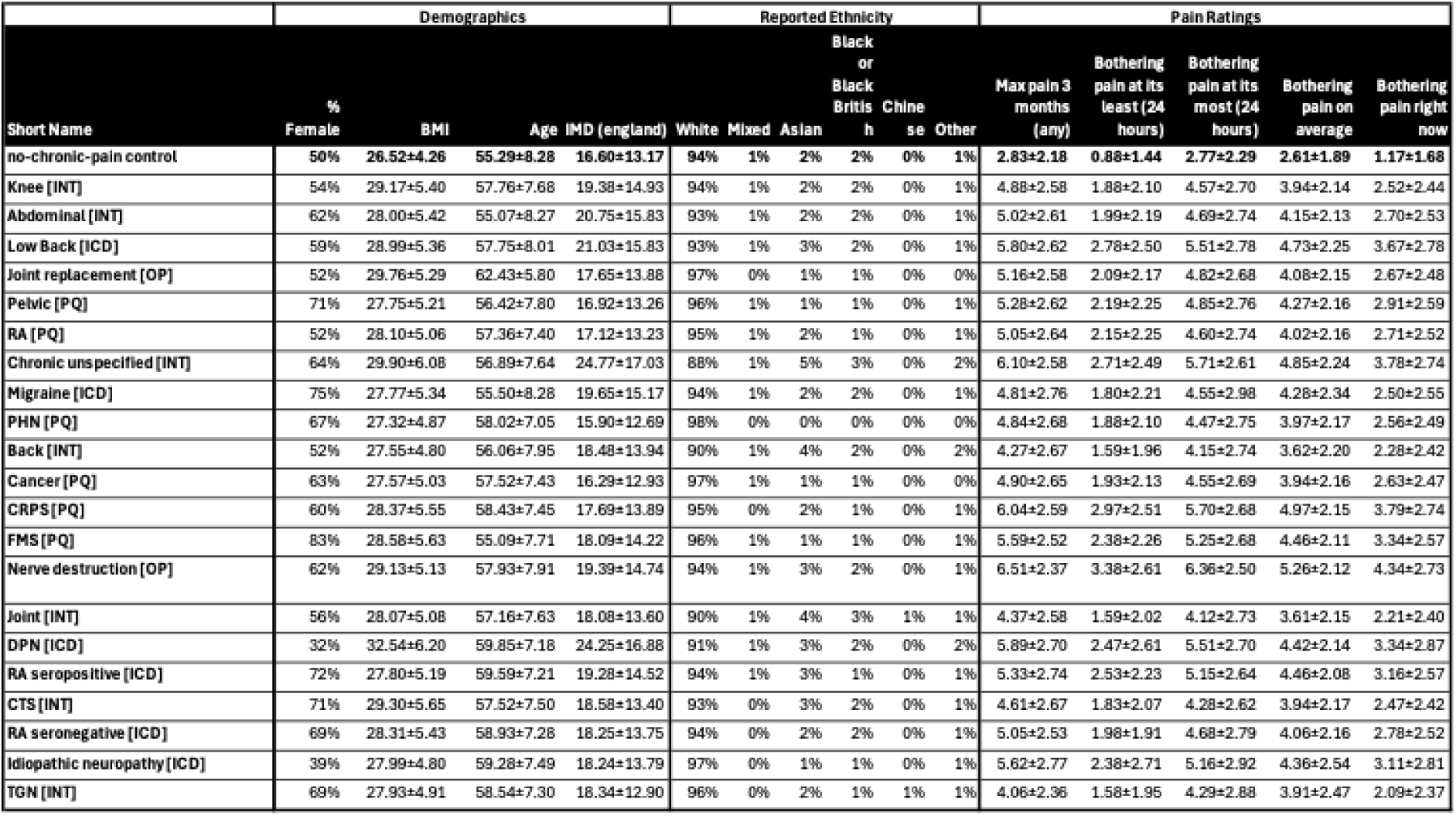
Demographic features.

To understand the comorbidities of the pain phenotypes the Jaccard index was calculated between each pair of phenotypes using the size of intersection divided by the total number of participants in the groups. To understand the demographics we summarized the age, sex, ethnicities, index of multiple deprivation and numeric pain rating scores within the pain questionnaire.

### Control cohorts

We defined a stringent ‘pain-free’ control cohort excluding participants with any chronic conditions which was used to compare with all pain derived phenotypes. Participants were included in the ‘pain-free’ control group if they had no pain related diagnosis (ICD9/ICD10/interview), no pain related procedure (OPCS4), no reported prescription of common chronic pain medications and no ongoing (interview/pain questionnaire) pain symptoms. Individuals were also excluded if no data were available in all the fields. Individuals were also excluded if no data were available in all the fields. For a full list of excluded ICD codes see supplementary table 1. This primary control group had 150,374 participants. To evaluate the efficacy of the optimised control approach, we employed a case-control GWAS in chronic knee pain to compare to the questionnaire result “Have you had knee pains for more than 3 months?” (field 3773) with cases as “yes” and controls as “no”. In addition, we used a frequently used larger control cohort of all other participants in the UK Biobank. The results were compared with previously published associations of knee pain in the UK Biobank [22]. A second alternative control cohort, comprising 235,483 individuals, was defined by excluding those who reported experiencing pain lasting more than three months in any of the following areas: knee (field 3773), hip (field 3414), stomach (field 3741), back (field 3571), neck (field 3404), face (field 4067), post-surgical pain (field 120005), or general chronic pain (field 2956). These control cohorts enabled us to investigate rare variant association studies across multiple phenotypes, endpoints, and continuous variables on a large scale[31].

### Principal component analysis

Because chronic pain is a pervasive issue that affects individuals across various ethnicities and populations worldwide, we included all ethnicities available in the UK Biobank and performed genetic associations in a diverse patient population.

To correct for this population structure within the UK Biobank we included principal components as covariates in our statistical models. The principal component vectors were calculated using the full PCA mode of Plink [32]. For each of the genotypic datasets (array and whole exome sequencing) we excluded the subset of variants in known inversions and MHC regions (chr6:25500000-33500000, chr8:8135000-12000000, and chr17:40900000-45000000) with a minor allele frequency of less than 1% and those within high linkage disequilibrium using the independent pairwise pruning, a 50kb window with steps of 5 as well as an r^2^ threshold of 0.2. After quality control and filtering steps we used 27051678 variants for our association analysis.

### Genetic association analysis

A generalised linear model regression using the allelic model was performed on the genotype array and whole exome sequencing data using plink2 [32] using the first 4 principal components, age and sex as covariates. We also performed calculations with 10 and more PCA with some cases in chronic knee pain which did not reveal significant differences for the genome-wide associations. Each of the pain cohorts were compared to the optimised pain-free control group described earlier. Variants with a missingness of greater than 5% in the UKB are not reported. We also calculated genetic associations based on quantitative measure like on the pain numeric rating scores from the pain questionnaire across different areas of the body (category 154). Gene level burden analysis was calculated using MAGMA [33] and variant associations that are corrected for linkage disequilibrium using the 1000Genome European reference cohort [34]. We applied a multiple testing correction that accounts for both the number of rare variants tested and the number of phenotypes analysed, using a standard genome-wide significance threshold across all analyses to ensure consistent control of type I error.

### Genetic annotations

To understand the impact of the variants, nominally significant variants (-log10(p)>5) were annotated for known pathogenicity using dbNSFP and predicted impact as well as gene annotations using SNPeff [35]. To find groups of related associations, we performed agglomerative clustering on the odds ratios of genome wide significant associations (-log10(p) >8). To investigate known associations with chronic pain, we annotated the associated variants with the human pain genetics database [14] and pain-relevant gene datasets (for a full list see supplementary table 2) from MSigDB v7.4 [36] joining by variant and gene annotation. In addition we annotated ion channels and GPCRs using the Uniprot database as they are critical to the function of nociceptors[16].

### Genetic correlation

To investigate genetic heritability and pleiotropy between chronic pain conditions we performed Latent Causal Variable analysis (LCV) [37] using the genotype array results. LCV analysis calculates a linkage disequilibrium adjusted genetic correlation between traits which it fits a latent variable that is causal for both traits. The ratio of the correlation of the latent variable to the two traits then determines the Genetic Causality Probability (GCP). A GCP score of 1 indicates that one trait is causal of the other (and –1 vice versa). As per the original authors recommendation we excluded variants within the MHC region and used linkage disequilibrium scores from the 1000G European reference panel and jack-knifed the calculation using 100 iterations.

### Epistasis

To generate testable hypotheses this research emphasizes analysis of rare variants, particularly those involving missense and loss-of-function (LoF) mutations, and we focus on specific conditions such as complex regional pain syndrome. We examined the epistatic interactions between a particular rare variant and a set of 1.5 million missense and LoF variants across the whole genome. This approach aims to better understand the penetrance of rare variants by genetic modifiers. Additionally, it helps elucidate the overall genetic architecture that may not be evident when analysing single genetic variants alone.

Consideration of epistatic interactions in the results from rare variant genetic associations can improve the accuracy and predictive power of genetic models for complex traits. For risk associations the epistasis was also run to identify potential protective variants. The calculations were performed using SNPint [38] using the logistic regression approach. Since epistasis calculations are computationally demanding, we reduced overall runtime by selecting variants with more than nine heterozygous carriers and medium or high impact as determined by SNPeff, and applied a multiple testing correction using a fixed significance threshold of (−log₁₀P>5.3), accounting for both the number of rare variants and the number of phenotypes analysed. [39].

## Results

### Demographic features of multiple pain phenotypes

There was a broad spread of demographic characteristics seen across the pain conditions. Some striking contrasts were evident in that there was a female preponderance for almost all the conditions (>70% for fibromyalgia, Migraine, Carpal tunnel syndrome and Pelvic pain) but a clear male predominance for diabetic painful neuropathy and idiopathic neuropathy (>60% male). The mean BMI across the pain conditions ranged from 27.32 in post herpetic neuralgia to 32.54 in diabetic polyneuropathy. Mean index of multiple deprivation score (IMD) from England, was lowest in General chronic pain of 15.9, and highest in diabetic polyneuropathy of 24.25. The estimated interquartile range (IQR) for deprivation scores was approximately 43.63 for individuals with general chronic pain and 43.64 for those with diabetic polyneuropathy, based on the observed mean scores and the full deprivation range from 0.53 to 87.8. Mean age of recruitment ranged from 55.07 in abdominal pain to 62.43 in joint replacement surgery (Joint replacement [OP]). The self-reported ethnicity was mostly white British ranging from 88.12% for general pain to 97.28% for joint replacement surgery. For demographic features of the cohorts see Table1.

The mean most bothering pain score ranged from 4.28 in carpal tunnel syndrome to 6.36 in “nerve destruction operation” (on a 0-10 scale). In all conditions, the median pain score for any single area was lower than the median of the highest pain reported across all areas. This highlights the differences in pain regionalisation of individuals within pain condition groups shown in Table 1.

Phenotype overlap within a phenotypic subset in pain endpoints can be quantitatively assessed using the Jaccard index, which measures the similarity between sets. By comparing the presence or absence of specific phenotypic traits across individuals or groups, the Jaccard index reveals the degree of shared characteristics. A higher index value indicates greater overlap, suggesting potential common biological mechanisms or shared genetic influences within the respective cohorts. This method is particularly useful for identifying clusters of pain phenotypes that may respond similarly to therapeutic interventions and a shared underlying pathophysiology (Table 2).

**Table 2:** Top Jaccard overlaps.

| Phenotype 1 | Phenotype 2 | Jaccard Index | N Unique Phenotype 1 | N Unique Phenotype 2 | Shared |
| --- | --- | --- | --- | --- | --- |
| **Low Back [ICD]** | Nerve destruction [OP] | 0.10 | 13,051 | 1,378 | 1,640 |
| **Knee [INT]** | Abdominal [INT] | 0.08 | 82,158 | 17,216 | 8,193 |
| **CRPS [PQ]** | Pelvic [PQ] | 0.07 | 2,681 | 10,239 | 995 |
| **RA seronegative [ICD]** | RA seropositive [ICD] | 0.07 | 915 | 1,041 | 148 |
| **RA [PQ]** | Pelvic [PQ] | 0.06 | 9,531 | 9,901 | 1,333 |
| **Back [INT]** | Joint [INT] | 0.06 | 4,485 | 2,282 | 453 |
| **PHN [PQ]** | Pelvic [PQ] | 0.06 | 5,374 | 10,220 | 1,014 |
| **FMS [PQ]** | CRPS [PQ] | 0.06 | 2,633 | 3,294 | 382 |
| **Chronic unspecified [INT]** | FMS [PQ] | 0.06 | 6,942 | 2,447 | 568 |
| **FMS [PQ]** | Pelvic [PQ] | 0.06 | 2,255 | 10,474 | 760 |

### Optimised control cohorts identify novel signals within knee pain

To investigate the effect of control cohorts we compared genetic associations for pain related conditions against different defined control groups using the genotypic array data (Figure 1). Our comparison of knee pain vs a non-specific control cohort replicates the previously noted association with rs143384 (-log10(p) 13.5, OR=0.94) in *GDF5* [22]. Using our control cohorts, we could also identify novel genome wide significant associations in *WDR46* (rs9277965 -log10(p) 8.1, OR=1.06),). These are expressed in pain relevant cell types like Schwann cells (*WDR46*), molecular mechanisms for neuronal differentiation (*TCF4*) as well as microtubule organization (*SDCCAG8*).

**Figure 1:**
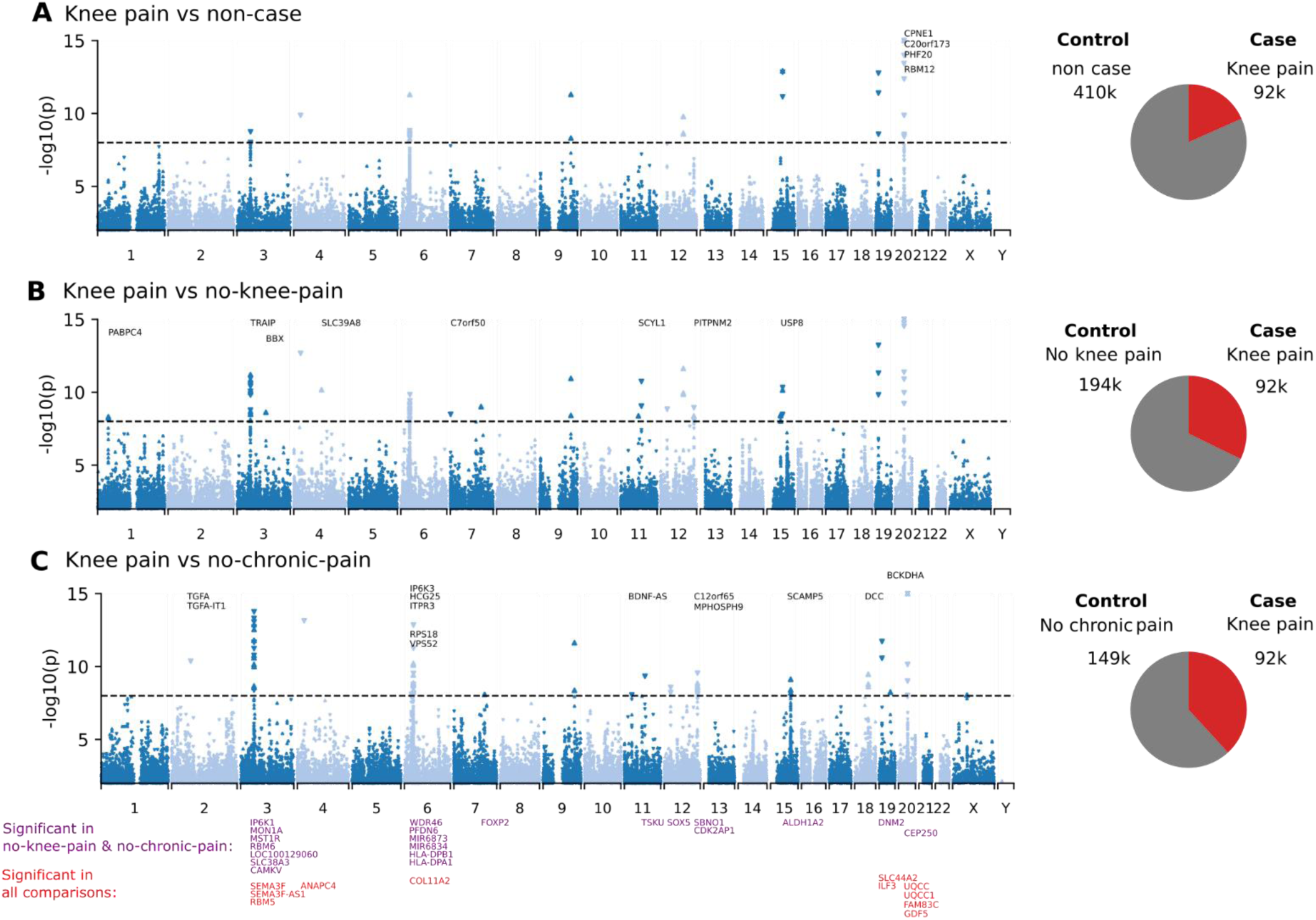
Genetic associations in array genotyping of knee pain with an active assertion of no knee pain (A), non-case (B) and excluding all incidence of chronic pain (no-chronic-pain) (C). Manhattan plots display the loci associations in each group with an upwards arrow for risk associations (odds-ratio greater than 1), and a downwards arrow for protective association (odds-ratio less than 1). The gene associations that are unique to that comparison annotated on each of the plots. The shared gene associations are displayed at the bottom of the figure, with those shared between all groups in red, and those shared between no-knee-pain and no-chronic-pain in purple. Pie charts on the right of each figure display the ratios of cases and controls.

**Figure 2:**
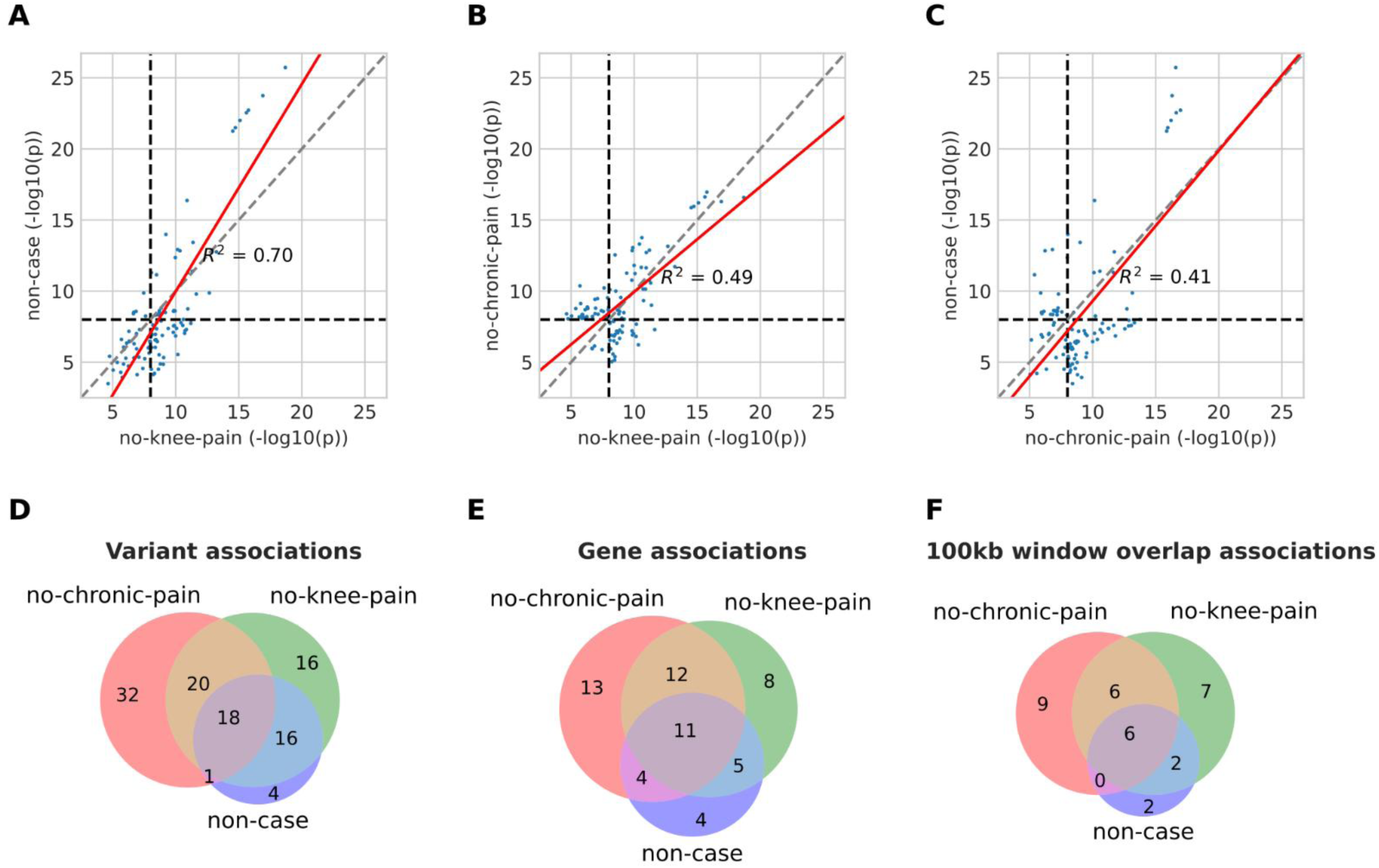
Summary of Genetic associations in array genotyping of knee pain with different control groups. All genome wide significant associations with -log10P >8 in at least one group compared pairwise (A-C). A dotted vertical and horizontal line at -log10P=8 and a diagonal line indicates a perfect correlation. Red solid line shows the linear fit between the pairs of association statistics. A summary of the difference in association significance between the No chronic pain control and a larger control cohort (D). Venn diagrams showing the number of overlaps in genes by any variant (E) and in 100kb windows (F).

**Figure 3:**
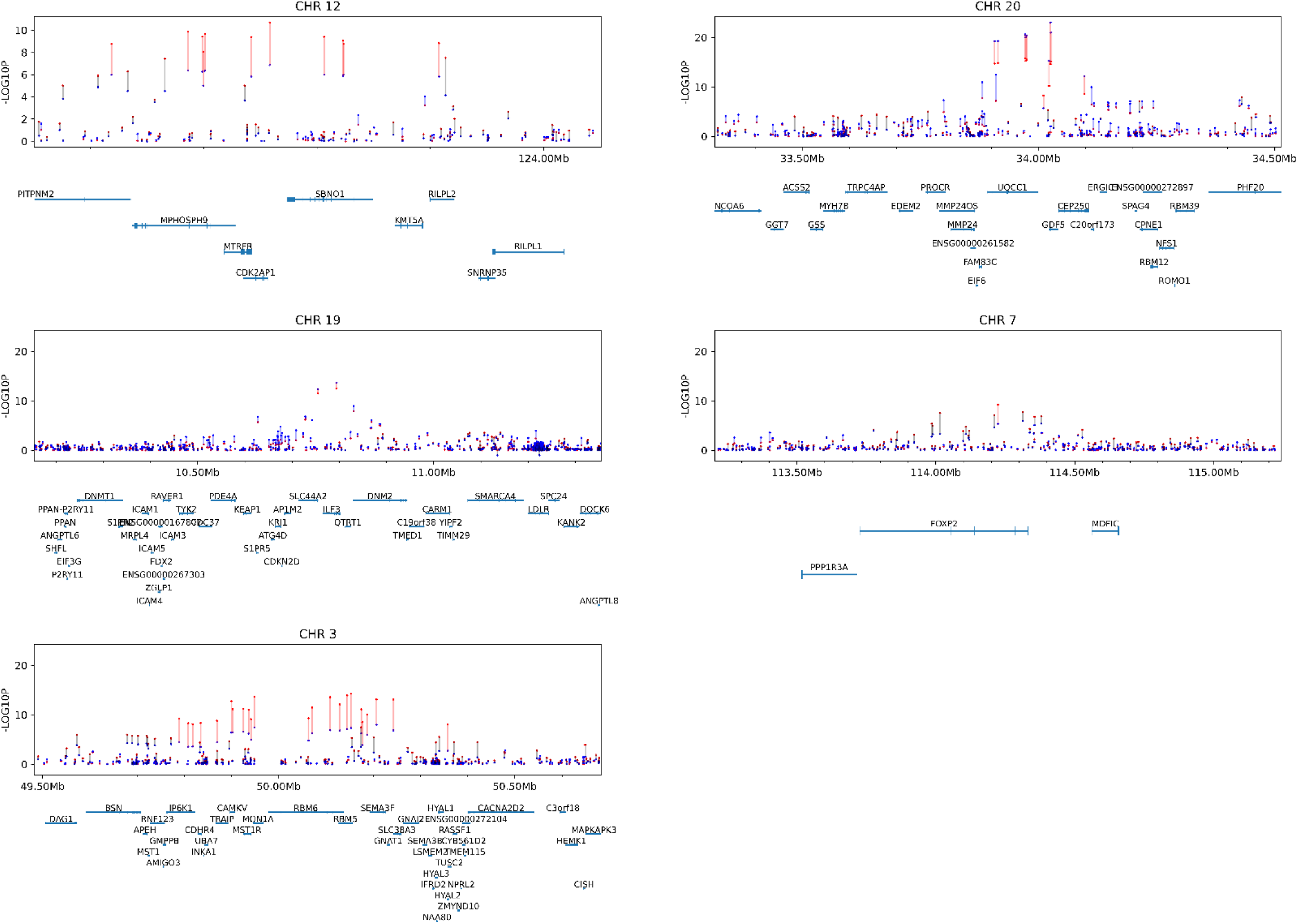
Fine mapping of the changes using optimized control cohort in chronic knee pain. Each point shows the -log10(p) for the association with UKB control (blue) or optimized pain cohort (red) with a line joining the two comparisons. A red or grey line indicates an increase in statistical association, with red showing a significant association in the optimized group. A blue line indicates a decreased association in the optimized analysis.

**Figure 4:**
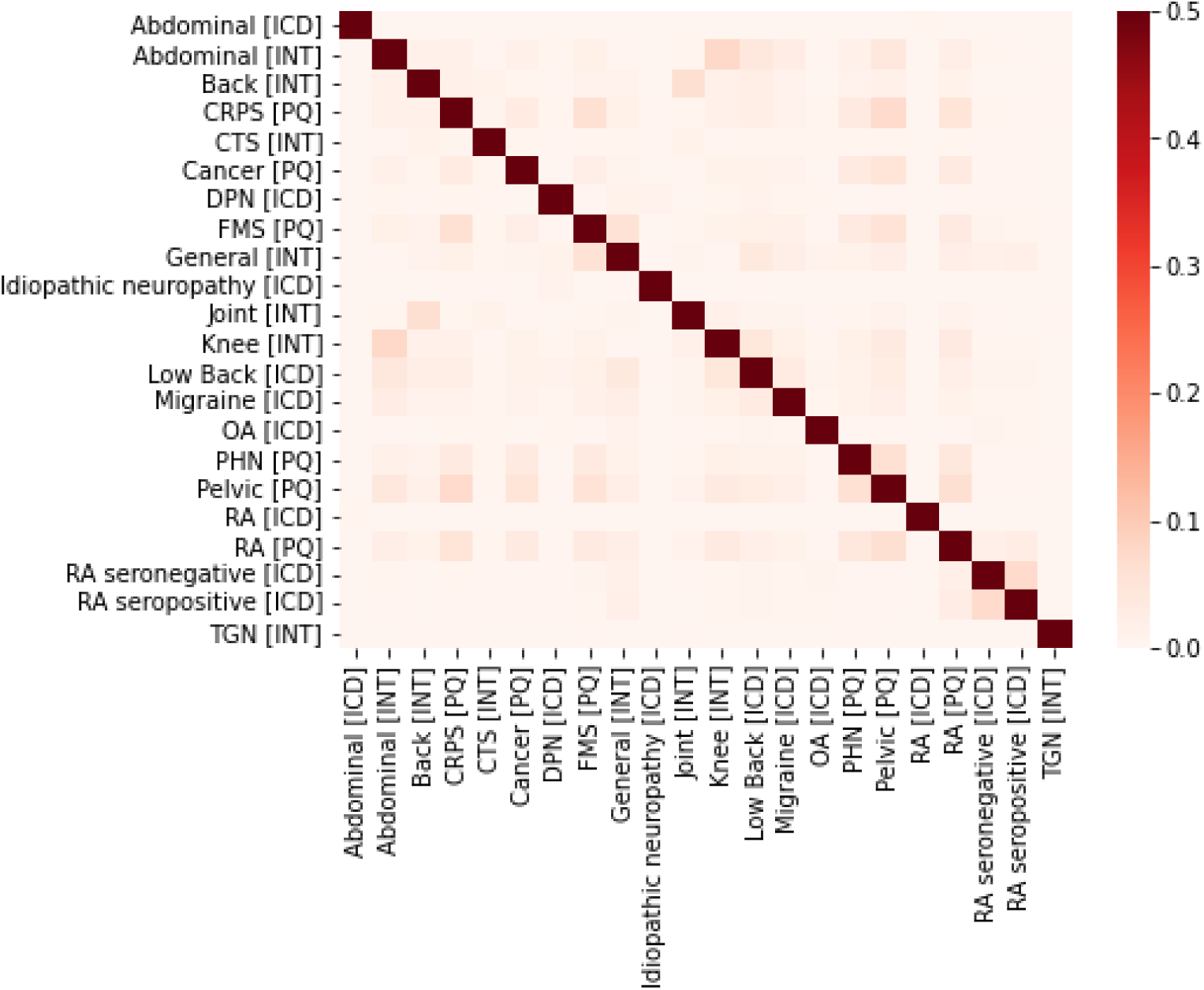
Jaccard index Phenotype overlap within phenotypic subset. A) Heatmap of the Jaccard index between the groups showing the intersection between the groups. Most phenotypes show low degree of overlap between the cohorts. See Table 2 for highest overlaps

**Figure 5:**
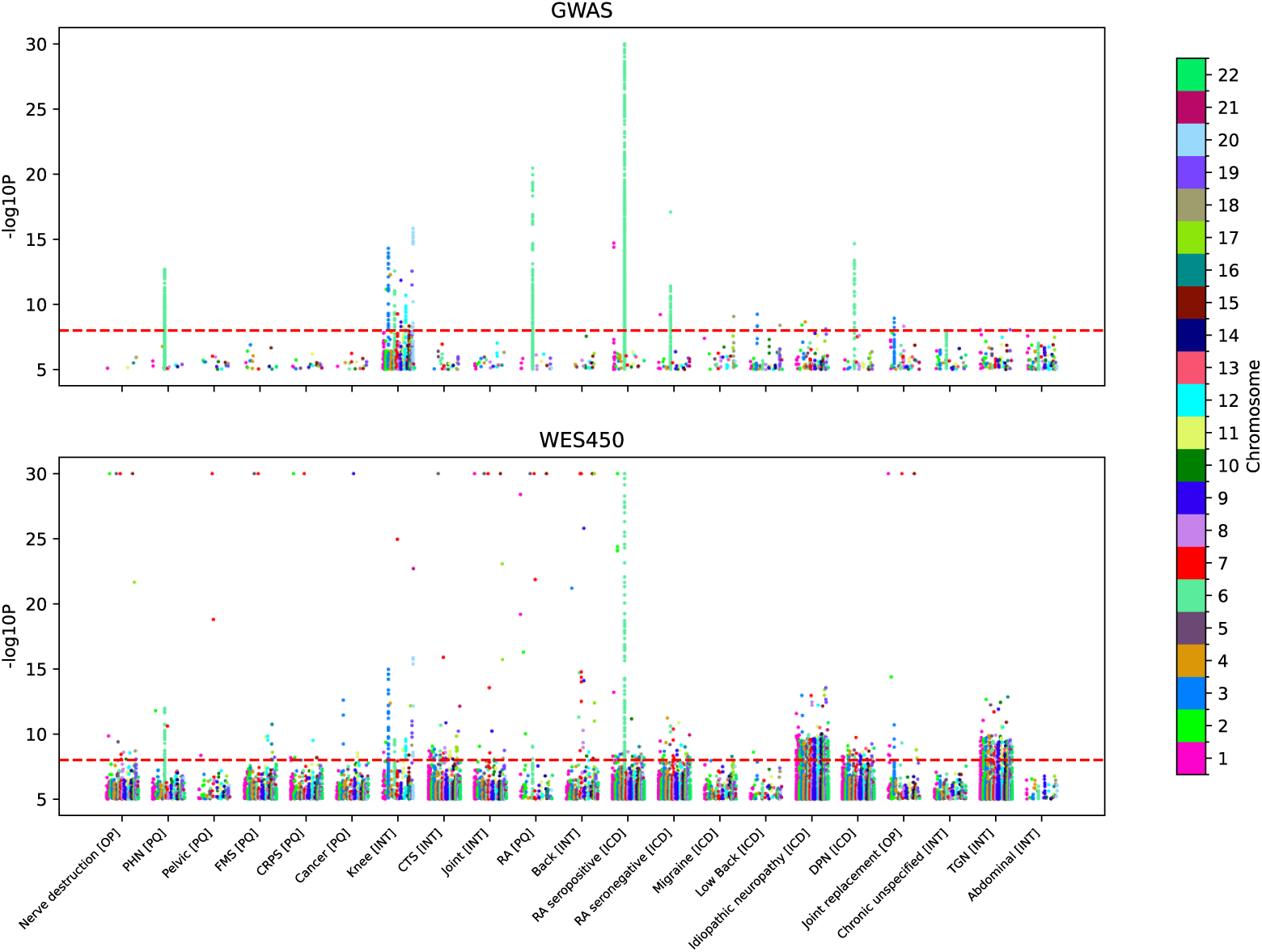
Variant level associations for chronic pain conditions across array and whole exome sequencing data using a common control in a subset of the results. Red line indicates a -log10(p) value of 8. Each point is a single variant association.

**Figure 6:**
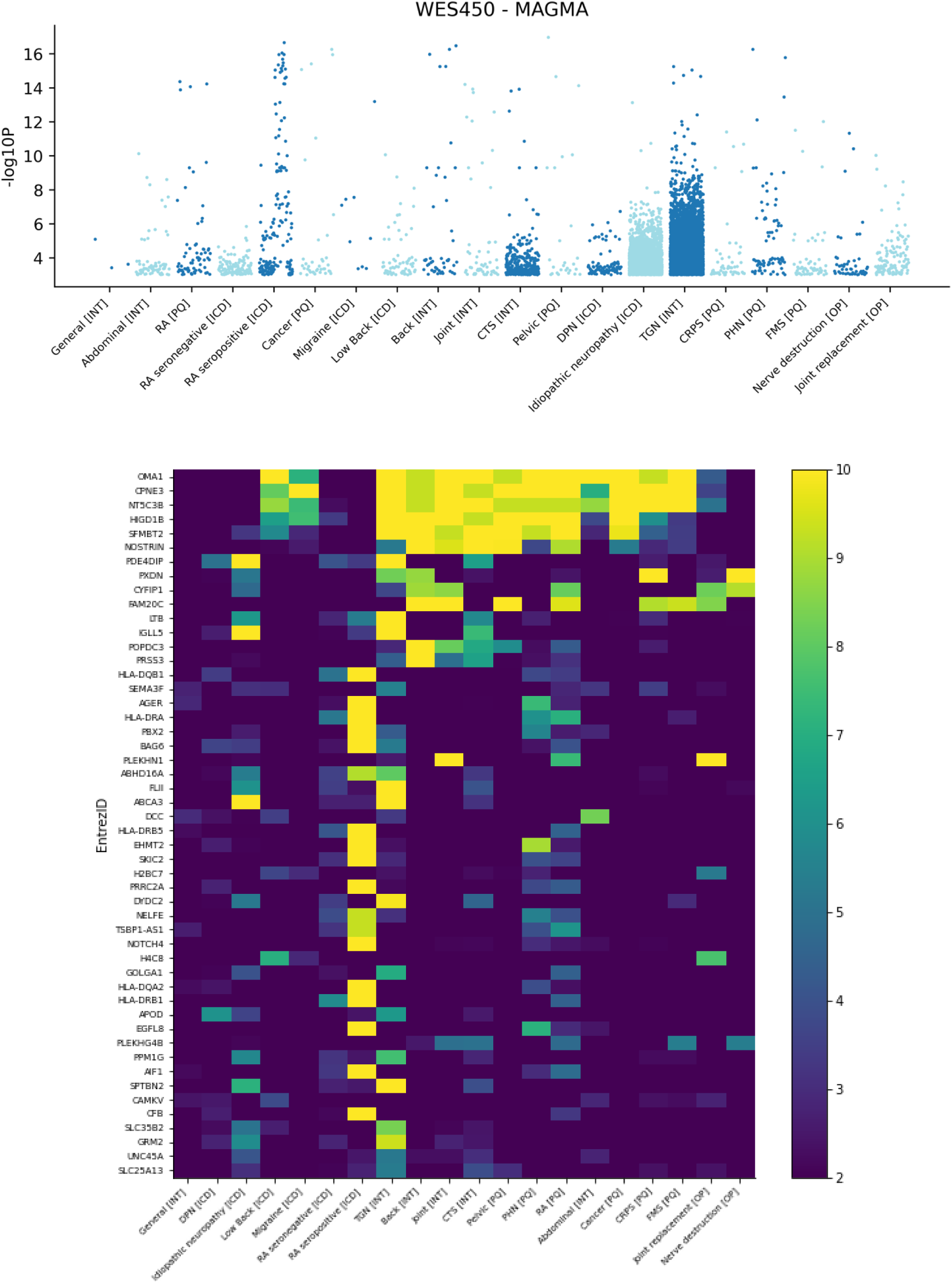
Gene level burden analysis for multiple chronic pain phenotypes. A) Manhattan. B) Heat map of genes. -log10(p) value in colour sorted by hits.

Within the whole exome sequencing associations and the optimised pain-free control cohort we found an increased significance within the *GDF5* locus, as well as at a locus on chromosome 3 for *RBM6/SEMA3F/CAMKV*, and in the HLA region on chromosome 6 which is relevant for any autoimmune disease. This optimised control analysis showed reduced variant associations on at *SLC44A2* locus on chromosome 19; and a novel association at *FOXP* locus on chromosome 7. Based on these encouraging results and the detection of novel associations we decided to use this pain-free control group for our associations as well as additional control cohorts to test the sensitivity of rare variant associations for some endpoints.

### Missense (non-synonymous) variant associations for chronic pain

The generalised linear model against the pain-free control cohort revealed many genome-wide significant (-log10(p) >8) associations within the chronic pain conditions that we have summarized in form of a tabular database. For the full list of associations see supplementary table 3. We examined the inflation of test statistics, which serves as a measure of false positives (type I error), across 329 chronic pain phenotypes. Our analysis identified 245 phenotypes with lambda values ranging from 0.8 to 1.2. Given the complexity of genetic associations in mixed populations and the presence of rare variants from whole exome sequencing, this range allows for some degree of inflation without indicating a significant problem with false positives, thus maintaining the integrity of our findings. Many of the previously reported variants and genes from previous GWAS of pain conditions could be confirmed (e.g. *SCN10A*, *GDF5*, *SOX5*) so we have especially focused on describing the novel variants which can be either found in specific chronic pain conditions but also those that showed on communalities across chronic pain conditions that are of particular interest for potential shared biological mechanisms and pathways (Table 3). In the following section we describe our novel findings for Fibromyalgia and CRPS in the context of rare variants in complex diseases (Table 4).

**Table 3:**
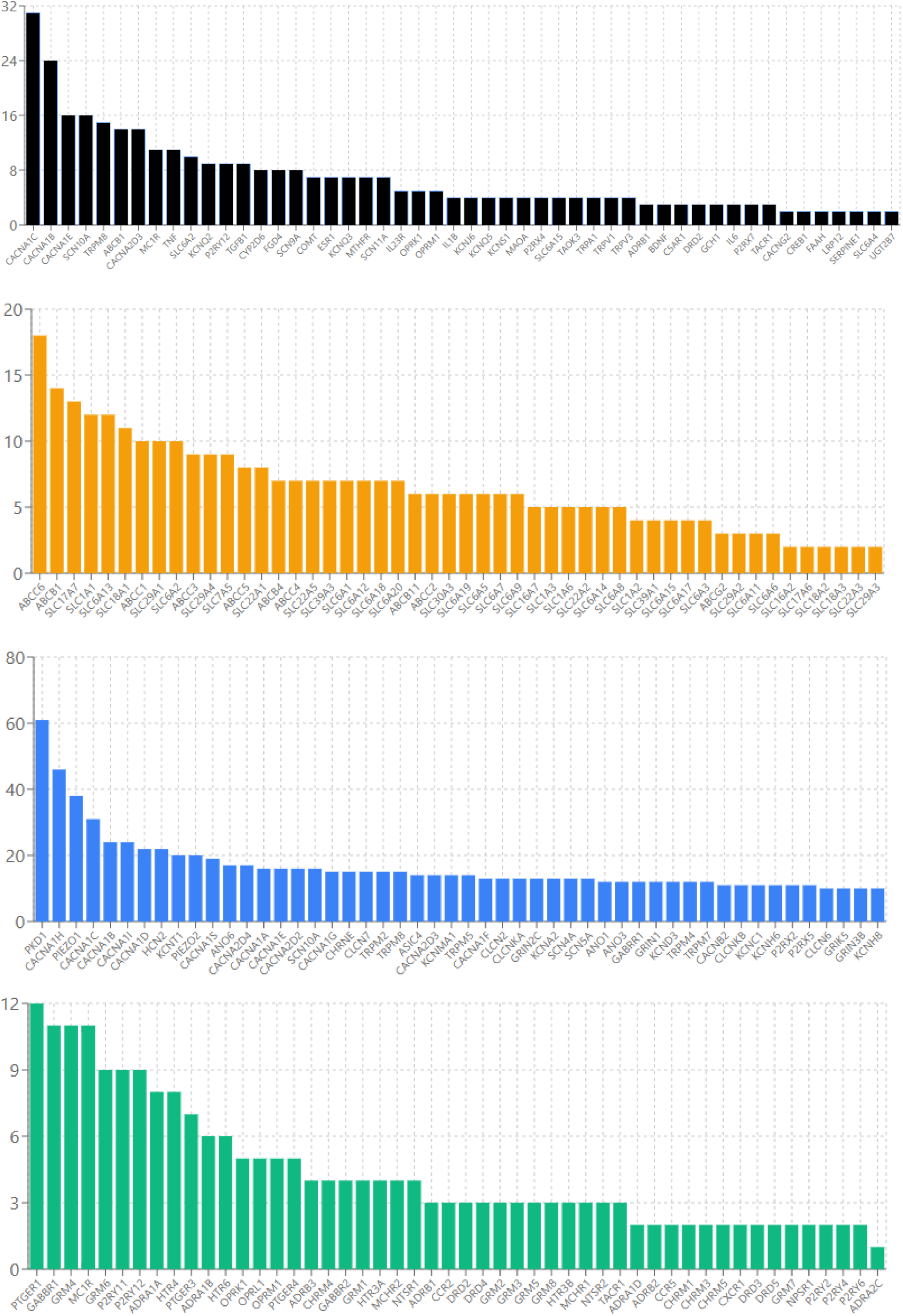
Common genetic associations shared across phenotypes.

**Table 4:**
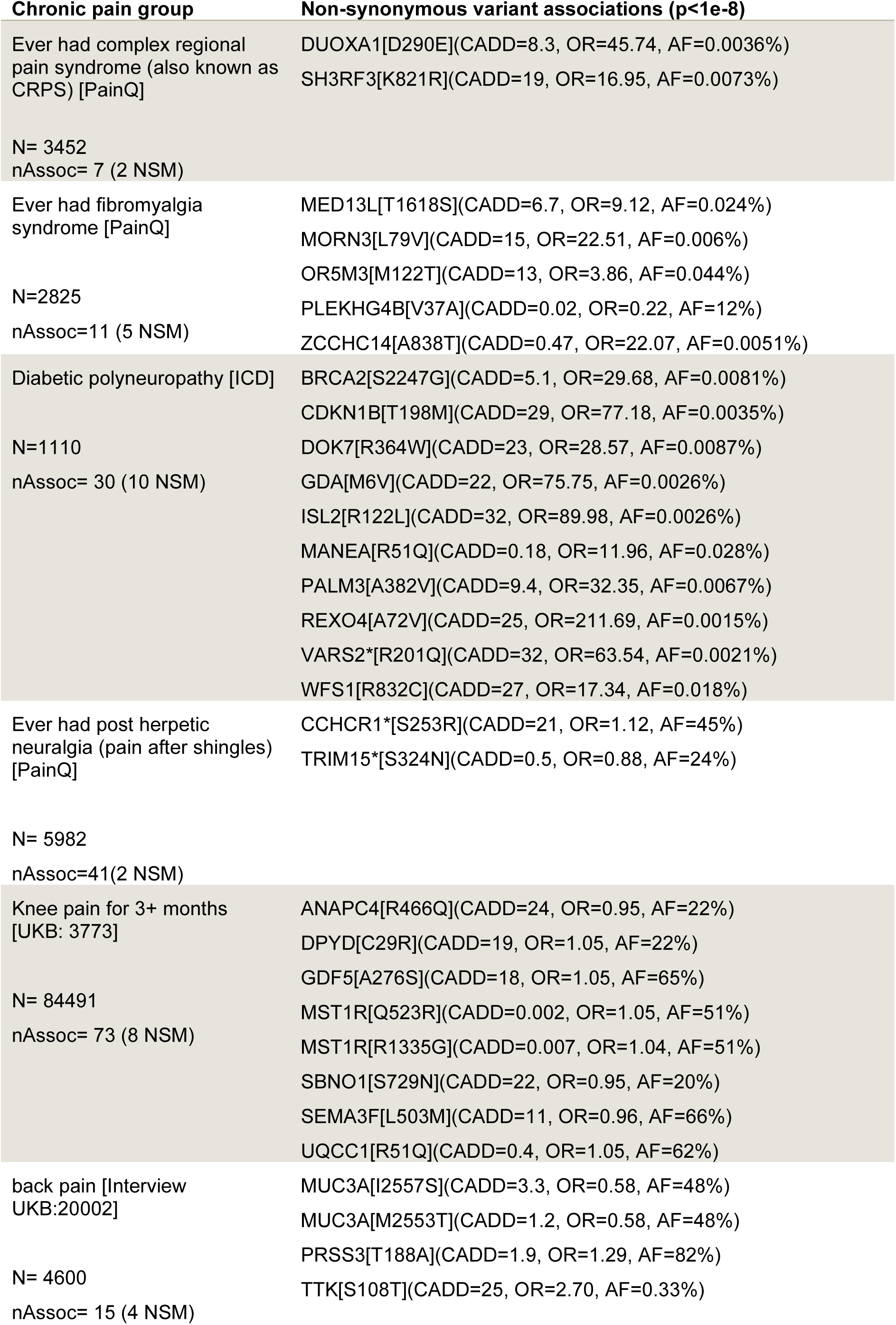
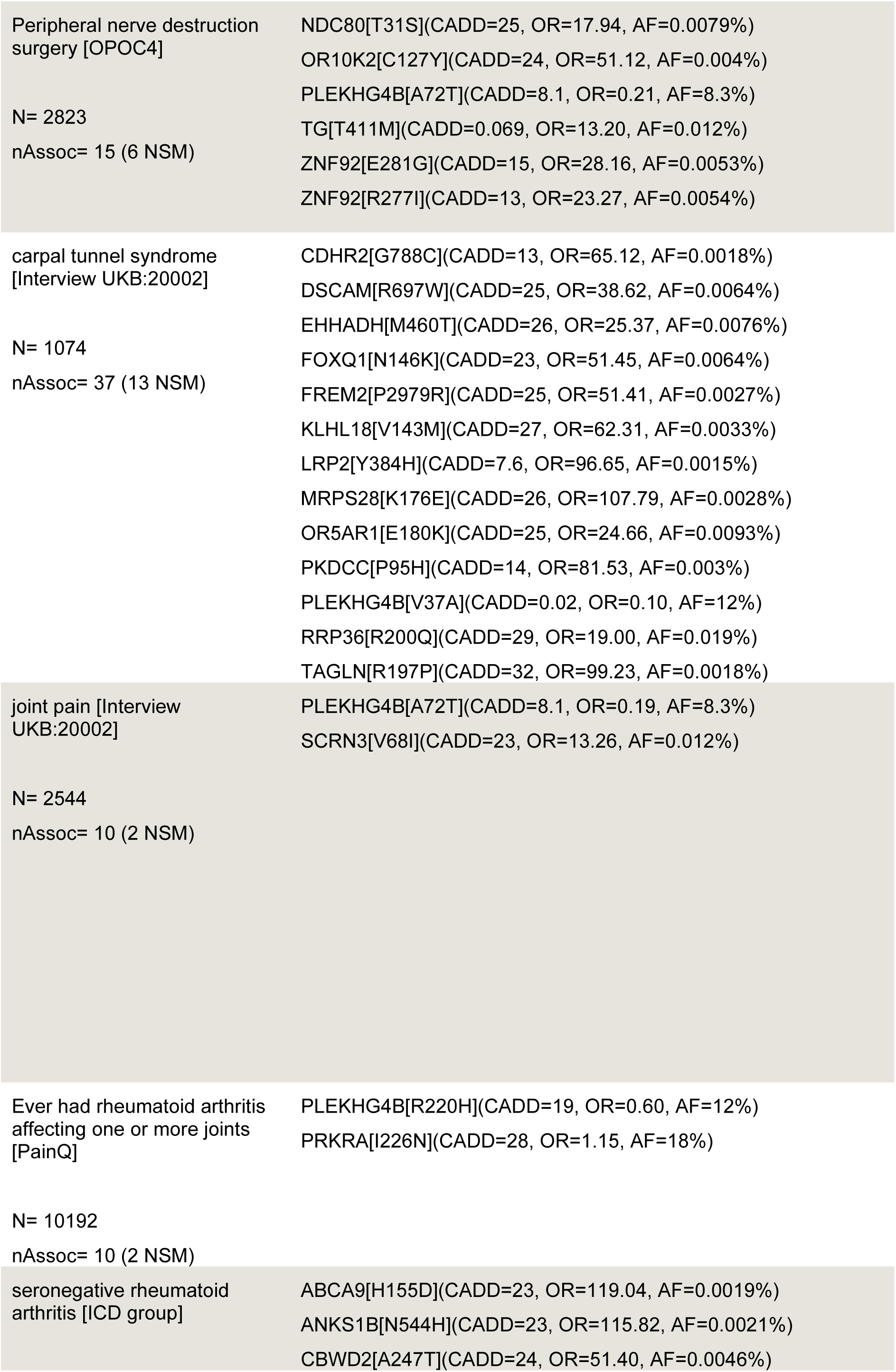
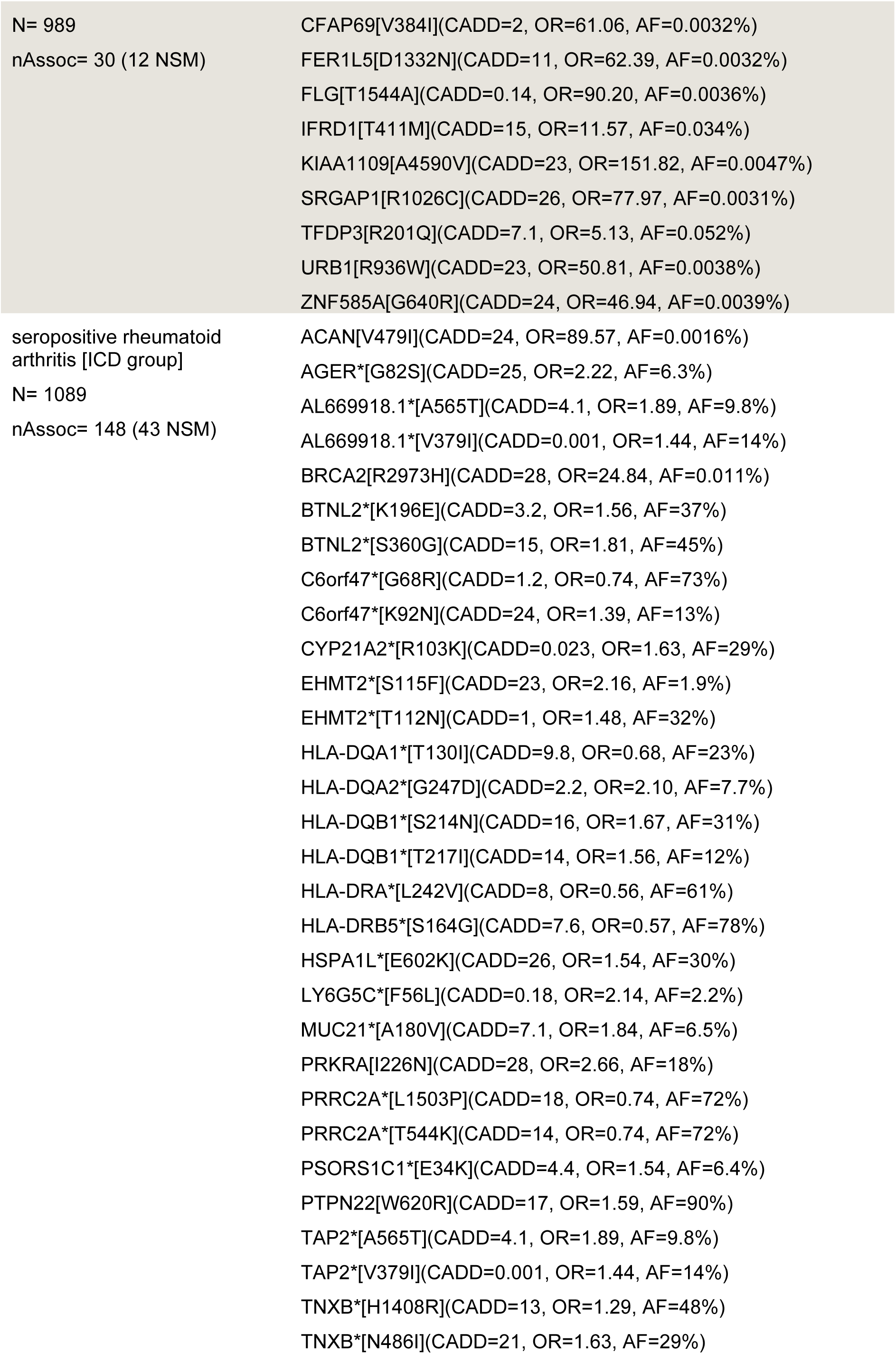
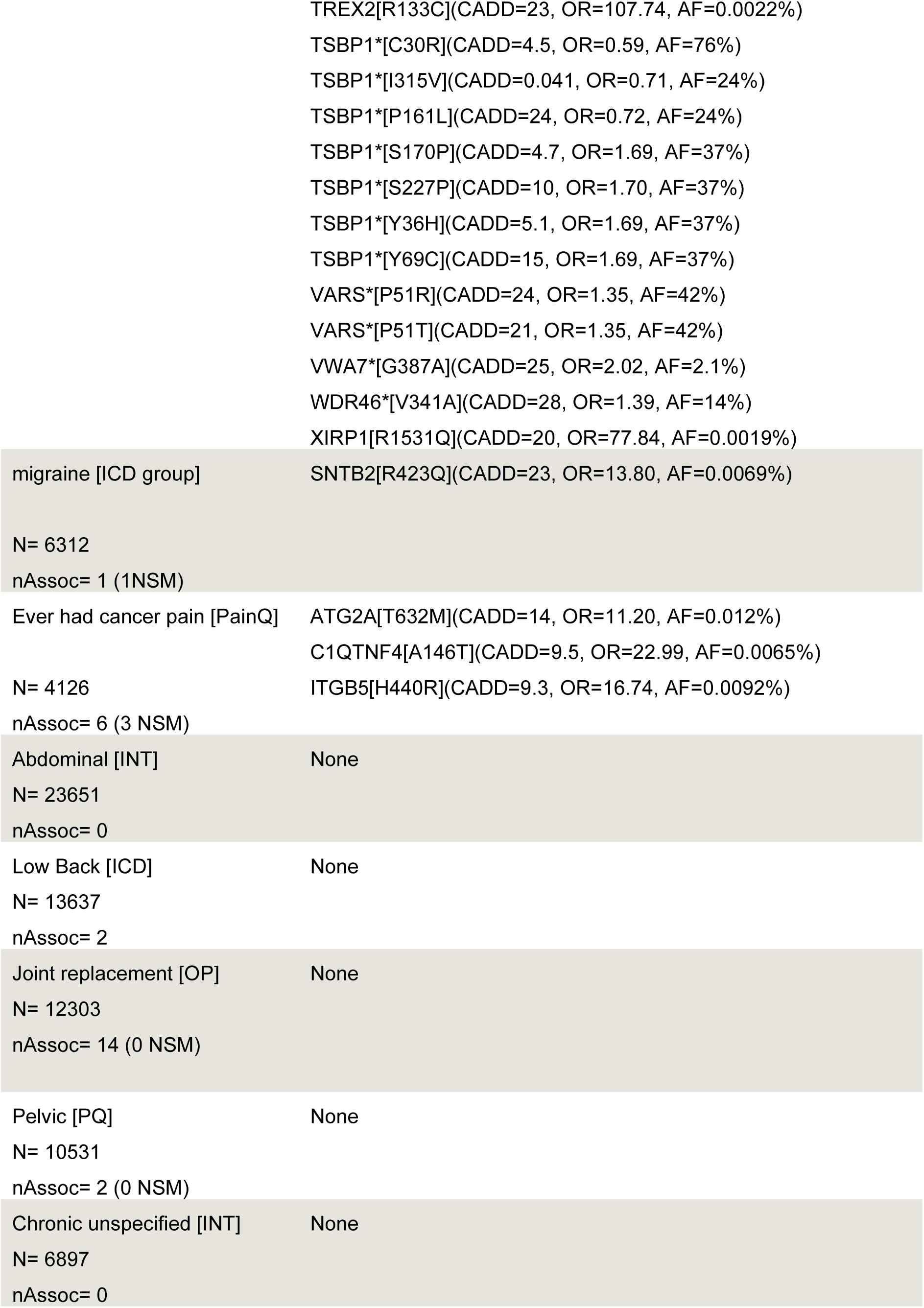
Unique non-synonymous genetic associations of chronic subset.

### Fibromyalgia

Fibromyalgia is a chronic widespread pain with muscular tenderness, often with fatigue and cognitive dysfunction [40]. In the general population fibromyalgia had a much higher incidence in females [41] which is replicated in the UK Biobank (4.9 fold higher in women). Previous GWAS studies have shown associations with *COMT*, *OPRM1* and *HTR1B* [42], which were not replicated in this study. This study revealed novel non-synonymous associations with Fibromyalgia of variants in *OR5M3*, *MORN3*, *MED13L*, *ZCCHC14*.

*OR5M3* is an olfactory related protein that associated with fibromyalgia uniquely across the pain conditions through a rare non-synonymous variant rs200070203 (M122T). Recent studies have implicated hypersensitivity to sensory stimuli, including olfaction within fibromyalgia [43]. While *OR5M3* may contribute to hypersensitivity, we found a specific variant rs200070203 that can be a driver to account for the observed effects since it is a rare variant with a low allele frequency. In our case 10 individuals with fibromyalgia carried at least one copy of the minor allele. *MORN3* associated with fibromyalgia through a rare non-synonymous variant rs750780231 (OR=22.51). There were 6 heterozygous carriers within the fibromyalgia cohort, and all were female. While this may be a low count, in the whole UK Biobank there are only 35 female heterozygous carriers of rs75078023 (total = 56), and 0 (total=13) within the pain-free control cohort. The role of *MORN3* is not well characterised but has been shown to interact with p53 leading to post-translational-modification and degradation [44]. *MORN3* is highly expressed in the brain, testis and fallopian tubes [45]. A recent single cell transcriptomic study described epithelial specificity to *MORN3* [46]. This finding may add strength to the link between female reproductive tissues and fibromyalgia, such as previous comorbidities with dysmenorrhea and endometriosis [47].

*ZCCHC14* was associated with fibromyalgia by a rare non-synonymous variant rs577390217 (A838T). There were only 5 heterozygous carriers with fibromyalgia, who were all female, 11 were in the control out of a total of 32 female carriers in the UK Biobank. *ZCCHC14* has been shown to be involved in antigen presentation in response to viral infection, specifically shown for hepatitis B [48]. There is an increased prevalence of fibromyalgia after hepatitis B infection [49] with a hereto unknown mechanism, and while this is a rare association it warrants further investigation of *ZCCHC14* and its interactors.

### Complex regional pain syndrome

Complex regional pain syndrome is a chronic pain condition that often occurs after injury or surgery where the pain is typically restricted to a single limb. The estimated prevalence of 20.57 per 100,000 person years [50, 51], In the UK Biobank there are 3,676 self-reported cases of complex regional pain syndrome which approximates to 12.9 per 100,000 person years when considering the baseline recruitment age for the full cohort.

For complex regional pain syndrome (UKBB data-field 120004) we found several genome wide significant associations using our extended control cohort such as a causal rare missense variant in sialic acetyl esterase (*SIAE* R479C, p-value 3.34E-08, odd ratio 7.1, MAF 1e-04) which has been reported to have a defective enzyme activity and known causal links to autoimmune diseases (e.g. rheumatoid arthritis) [52, 53]. Further analysis of heterozygous carriers of rs200070203 revealed that 61% of those diagnosed with CRPS were female, among a total of 57 carriers. This sex-specific distribution aligns with the established higher prevalence of CRPS in females, suggesting a potential gender-linked genetic susceptibility. Within the pain questionnaire 16% reported having complex regional pain syndrome whereas only 2% in our control cohort. Since we are focusing in this analysis on protective rare missense variants, we found I480M (rs61749251) in Mammary Analogous Zinc Finger 2 (*MAML2*, p-value 6.57E-06, odd ratio 0.62, MAF 0.015) which acts as a transcriptional cofactor for Notch proteins. The absence of *NOTCH* proteins in adult vessels leads to a higher endothelial proliferation and a reduced junctional integrity [54]. Together with mutations found in epistasis where *JCAD* regulates blood vessel endothelial cell proliferation, the gene *PLA2G7*, a lipoprotein-associated calcium-independent phospholipase A2, plays a pivotal role in the catabolism of phospholipids, particularly under conditions of inflammation and oxidative stress. *PLA2G7* effectively prevents the pathological accumulation of oxylipins within the vascular wall which is a process implicated in vascular dysfunction and chronic pain syndromes [55].

Our findings reinforce the concept that vascular pathophysiology is not merely a secondary consequence but a central driver of disease progression and pain perception. Genetic evidence from our UK Biobank analyses highlights a protective rare variant in *PLA2G7* (e.g., S388P) that is associated with reduced incidence of complex regional pain syndrome and fibromyalgia, suggesting a mechanistic link between lipid metabolism, vascular integrity, and nociceptive signalling. Overall, this supports a broader framework in which blood vessel dysfunction that mediated by inflammatory lipid species and endothelial stress responses that serves as a key modulator of chronic pain states. Targeting *PLA2G7* and related vascular pathways may thus offer promising avenues for therapeutic intervention in pain-related disorders.

Previous common variant based studies of complex regional pain syndrome did not show any significant associations beyond the HLA region[56] which were not replicated in subsequent studies [12], whereas small cohort studies have identified variants in *ANO10*, *P2RX7*, *PRKAG1* and *SLC12A9* [57] [58]. In this analysis, we saw no indication of rare variants in the HLA region in CRPS, nor any of the variants identified in the smaller cohort studies. We did identify novel associations including non-synonymous variants in *SH3RF3* (K821R), an E3 ligase involved in ubiquitination[59]; *NT5C3B* (S213C; A209V), which hydrolyses methylated Guanosine nucleotides (specifically m7G) [60] and *DUOXA1* (D290E) which is required for hydrogen peroxide synthesis in the thyroid. How general cellular processes such as in *SH3RF3*, and *NT5C3B* influence the risk of CRPS may be difficult to interpret; However, thyroid dysfunction has previously been associated with neuropathic pain [61] and perhaps this non-synonymous variants in *DUOXA1* may influence susceptibility. Further studies on the functional consequence of D290E on *DUOXA1* is needed to test this hypothesis.

We identified a high genetic causal probability of chronic pain on complex regional pain syndrome suggesting that the genetics of chronic pain are sufficient to explain the shared heritability between the conditions. It may be that environmental factors or other higher impact rare variants underly the development of complex regional pain syndrome in individuals already genetically susceptible to chronic pain. In partial support of this familial cases of CRPS typically have an earlier age of onset and increased severity [62].

We further analysed our genome-wide significant association in sialic acetyl esterase (rs376857712, *SIAE* R479C, p-value 3.34E-08, odd ratio 7.1, MAF 1e-04) for epistatic interactions with a genome wide set of 1.5M missense variants. Such epistatic interactions can modify the penetrance of pathogenic mutations and to identify additional functional effects from these mutations and their biology. In this case we found the mutation *V366E* in *JCAD* (beta 3.4, CHISQ 20.4, OR 29.9, p-value 6.39E-06) reported to play a key role in coronary artery disease which might link to the related blood vessel pathophysiology. While the identified variant in *PLA2G7* (S388P) does not reach conventional genome-wide significance (p = 3.16×10⁻⁴), its biological relevance remains compelling. The variant demonstrates a strong protective effect (OR = 0.12) and is supported by a robust chi-square statistic (CHISQ = 12.9), suggesting a meaningful association. Importantly, *PLA2G7* plays a critical role in phospholipid catabolism during inflammatory and oxidative stress responses, mechanisms central to vascular pathophysiology [63, 64]. The observed protection against oxylipin accumulation in the vascular wall aligns with known functions of PLA2G7 and supports its potential contribution to disease modulation.

### Novel pain loci across multiple pain phenotypes

To identify common molecular mechanisms, we analysed the most frequently associated variants across multiple chronic pain phenotypes and found several genetic loci like *PLEKHG4B* and *PRKRA*. For *PLEKHG4B* we identified 9 associated variants including rs12516846, rs12523402 and rs13436090 that have a protective effects (odds ratio < 1) in Rheumatoid arthritis and Fibromyalgia. *PLEKHG4B* is a guanine nucleotide exchange factor which plays a role in the regulation of the cytoskeleton. In the context of autoimmune diseases, it can affect the activation and migration of immune cells and lead to a dysregulation of immune responses that are often observed in these conditions [65]. Further, cytoskeletal dynamics are closely related to neuronal outgrowth and sprouting where actin filaments, microtubules, and intermediate filaments, which provide structural support for the growth and guidance of neuronal processes. Herein, actin filaments are important for the growth of neuronal cones, which drive the extension or retraction of neurites leading to the plasticity of the nervous system [66]. *PRKRA* had 6 variants with genome wide significant associations rs75862065, rs77419724, rs3997876, rs62176107, rs62176113 and rs9406386. Of these, rs75862065 was the most common association across conditions including low back pain, knee, joint and pelvic pain, seropositive rheumatoid arthritis, post-herpetic neuralgia and trigeminal neuralgia. It is an interferon-induced protein kinase that is central to the antiviral response, integrative stress response (ISR) response, and apoptosis. In the context of neurodegenerative diseases *PRKRA* modulates pro-inflammatory pathways via interaction with PKR and induces pro-inflammatory cytokines[67].

### Genetic correlation and known associations

We found high correlations in common variants across all pain conditions with a rho estimate ranging from 0.34 between Cancer pain and diabetic peripheral neuropathy to 0.90 between migraine and complex regional pain syndrome. This coefficient is commonly used to denote the genetic correlation as a measure of shared genetic architecture between two traits and can range from 1 to -1. It also measures the extent to which genetic factors influencing one trait are shared with those influencing another, offering insights into pleiotropy and common underlying biological mechanisms. For instance, indicating a role of central sensitization where the central nervous system becomes hypersensitive to stimuli, leading to an exaggerated pain response, neuroinflammation and autonomic nervous system dysfunction leading to symptoms such as changes in blood flow and temperature regulation [68].

Since we performed latent causal variable (LCV) analysis we also get the genetic causality proportion (GCP) that distinguishes genetic correlation from genetic causation between two traits. A GCP of 0.83 between chronic unspecified pain and complex regional pain syndrome indicates strong genetic causality, suggesting that the former is likely genetically upstream of the latter. This implies vertical pleiotropy, where genetic variants influencing chronic unspecified pain may directly contribute to the development of CRPS. In comparison with Fibromyalgia, we hypothesize that the autoimmune related genetic background might explain a common pathophysiology in between both diseases (for further associations see Table 5 and Figure 7).

**Figure 7:**
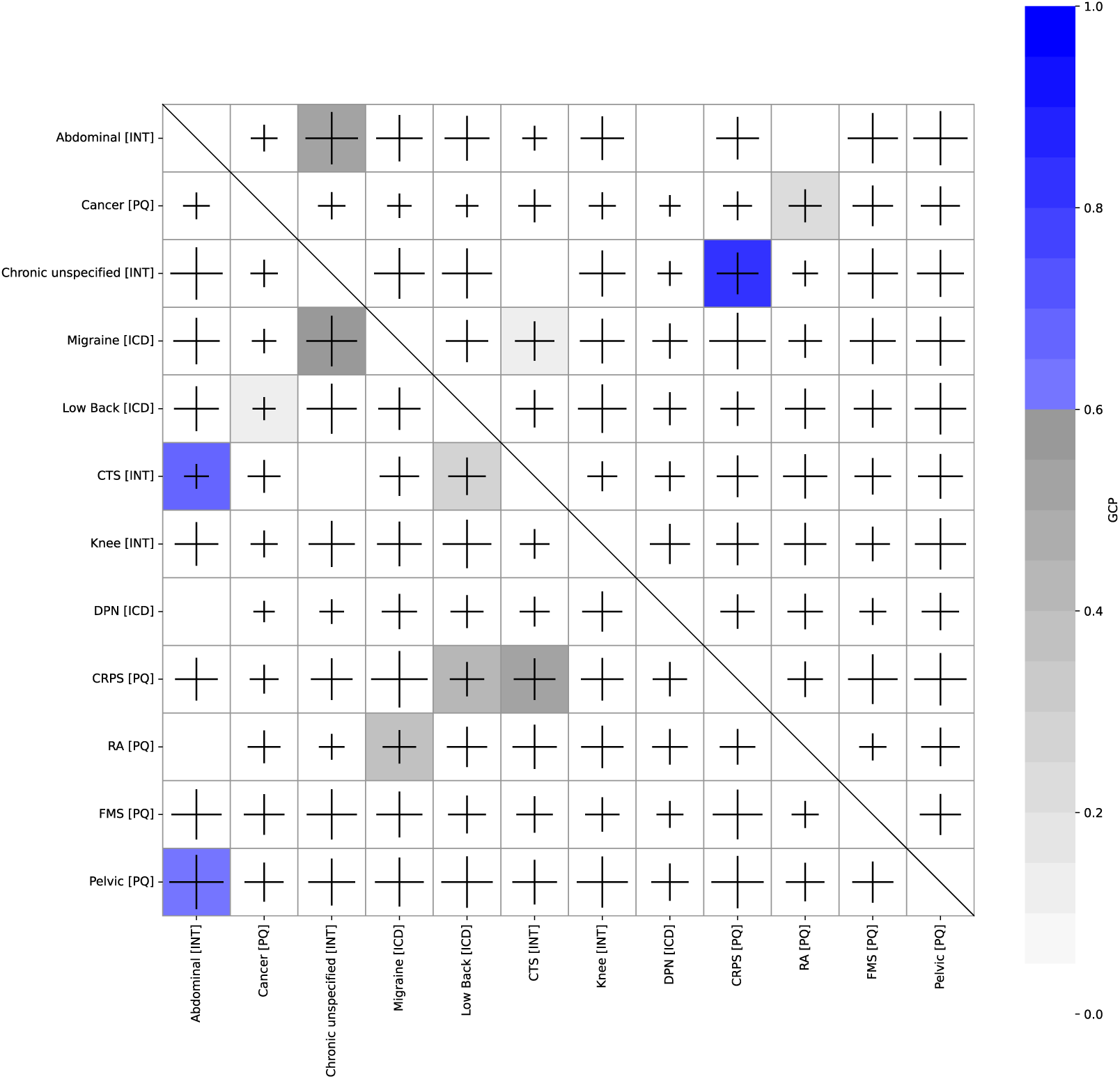
Shared Genetic architecture of chronic pain – results from latent causal variable analysis. The colour of each square indicates the GCP values where heritability is greater than 7 and GCP is significant (p < 0.05) for trait on row on trait on column. Blue colour indicates GCP over causal threshold >0.6, and grey a GCP value as suggestive. Significant genetic correlation is indicated with “+” for a positive genetic correlation between the traits, with the text scaled to the rho estimate. Empty squares highlight a lack of significant correlation between the traits.

**Figure 8:**
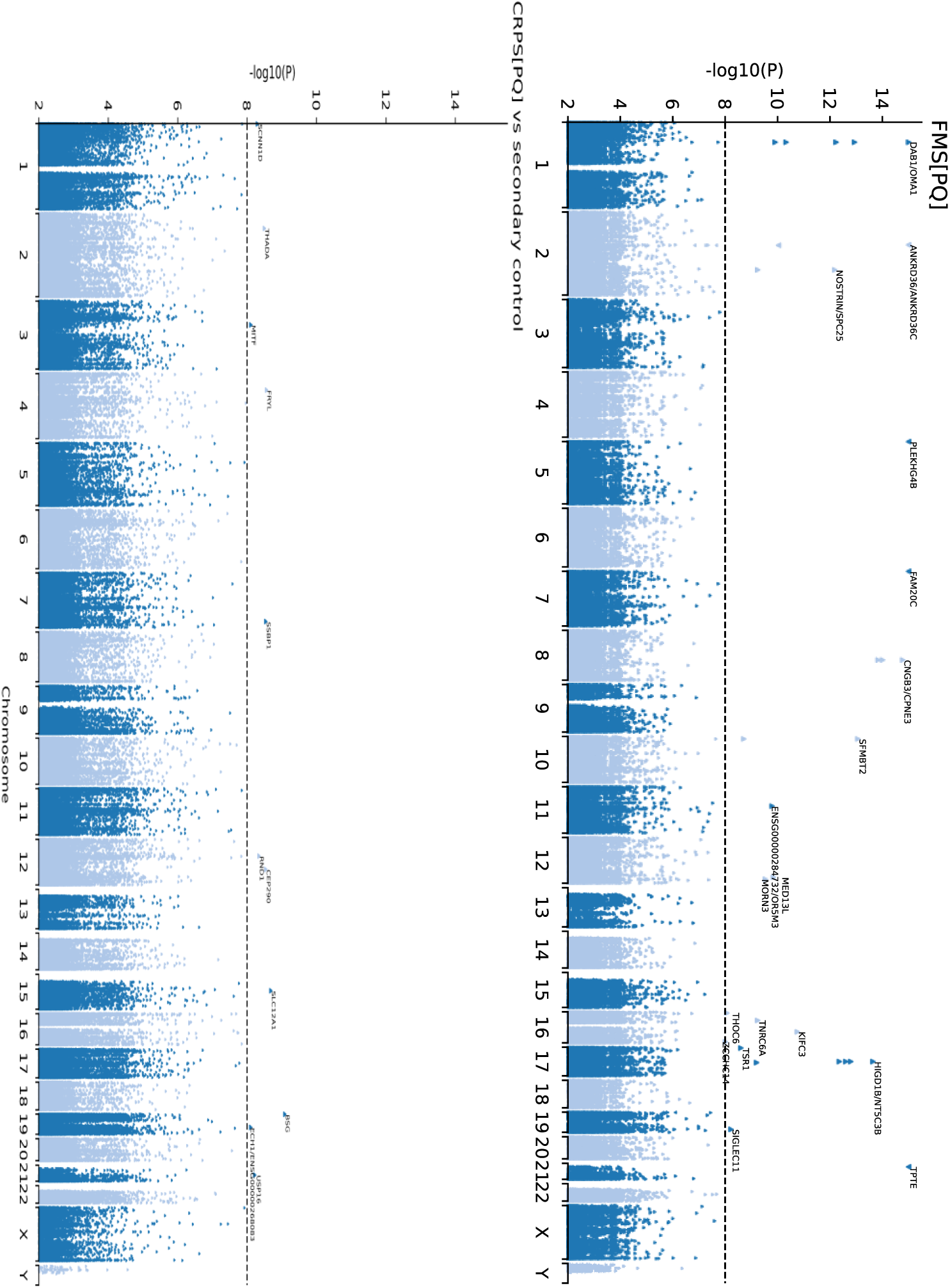
Manhattan plots for complex regional pain syndrome and fibromyalgia

**Table 5:**
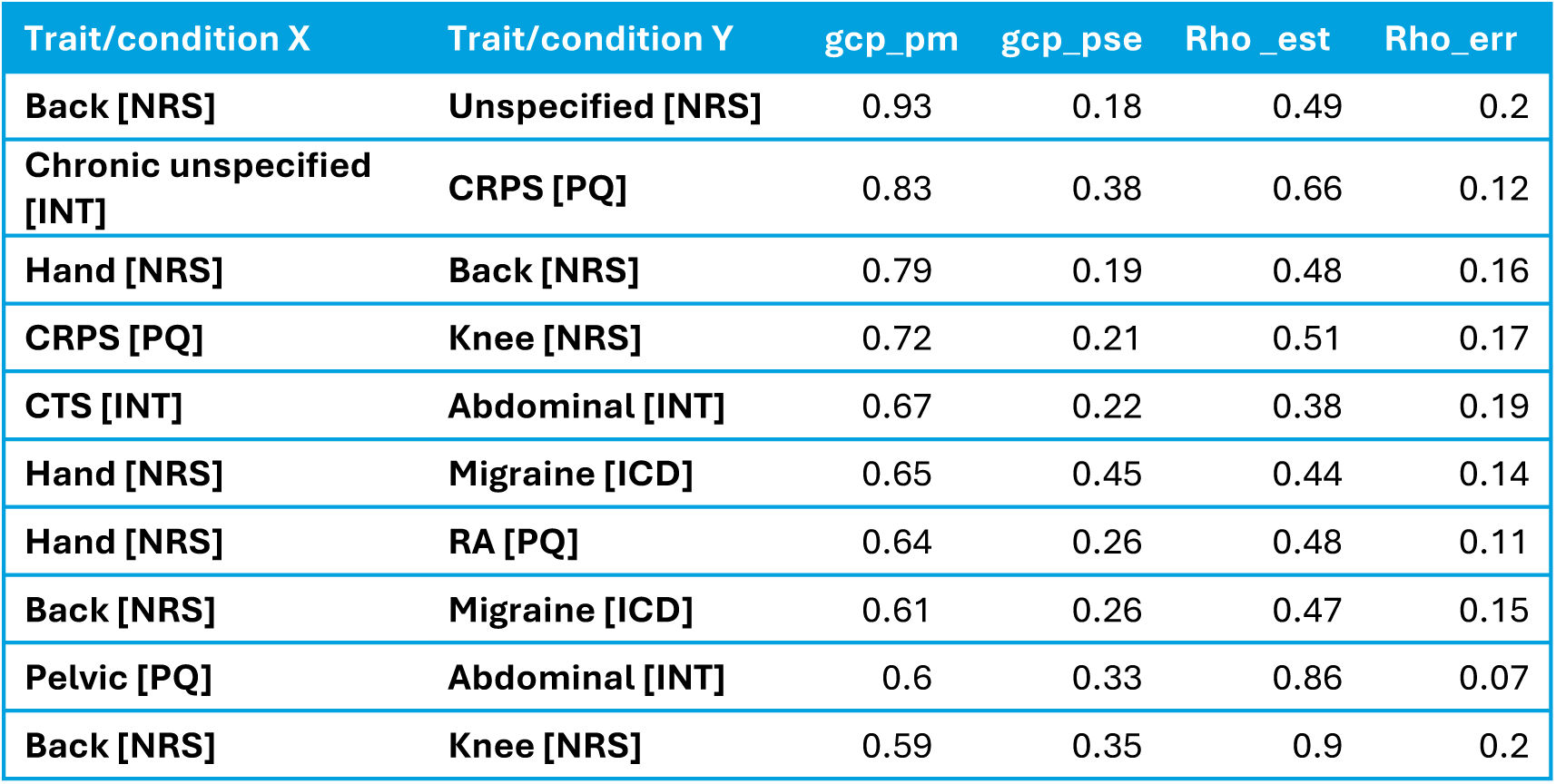
Genetic correlations.

| Trait/condition X | Trait/condition Y | gcp_pm | gcp_pse | Rho_est | Rho_err |
| --- | --- | --- | --- | --- | --- |
| Back [NRS] | Unspecified [NRS] | 0.93 | 0.18 | 0.49 | 0.2 |
| Chronic unspecified [INT] | CRPS [PQ] | 0.83 | 0.38 | 0.66 | 0.12 |
| Hand [NRS] | Back [NRS] | 0.79 | 0.19 | 0.48 | 0.16 |
| CRPS [PQ] | Knee [NRS] | 0.72 | 0.21 | 0.51 | 0.17 |
| CTS [INT] | Abdominal [INT] | 0.67 | 0.22 | 0.38 | 0.19 |
| Hand [NRS] | Migraine [ICD] | 0.65 | 0.45 | 0.44 | 0.14 |
| Hand [NRS] | RA [PQ] | 0.64 | 0.26 | 0.48 | 0.11 |
| Back [NRS] | Migraine [ICD] | 0.61 | 0.26 | 0.47 | 0.15 |
| Pelvic [PQ] | Abdominal [INT] | 0.6 | 0.33 | 0.86 | 0.07 |
| Back [NRS] | Knee [NRS] | 0.59 | 0.35 | 0.9 | 0.2 |

The human pain genetics database contains 537 unique genes, of which 304 associated with at least one pain condition in this analysis of the UK Biobank. Of these 304 gene associations 39 genes were in similar categories, 26 of which were with arthritis related conditions. See Supplementary table 3 for details of the overlapping conditions.

### Novel genes in pain target classes of GPCRs and ion channels

We have also investigated novel genes in classical target classes for pain, like G-protein-coupled receptors (GPCRs) and ion channels (Table 6). GPCRs modulate pain through their interaction with endogenous ligands such as neurotransmitters, endorphins and hormones, and mutations in either the receptor or endogenous ligand can enhance or inhibit pain signals[69] like reported for the COMT val158met polymorphism which has been linked to pain severity in Parkinson’s disease patients [70]. In our study we have identified several novel genes for peripheral neuropathy like hereditary motor and sensory neuropathy (ICD-10 G60), which is a group of inherited disorders that affect the peripheral nerves and is characterized by progressive loss of muscle tissue and touch sensation across various parts of the body [71]. The condition is genetically heterogeneous, meaning it can be caused by mutations in different genes. We have identified several new GPCRs like CELSR2/3, the neurotensin receptor NTSR2, ADGRA3 and MTNR1B with genome-wide significant associations and high odd ratios. CELSR2/3 receptors are members of the cadherin superfamily, which are involved in various cellular processes including cell adhesion, signalling, tissue morphogenesis and are of particular importance in the context of neural development [72]. ADGRA3 that is also known as GPR125 is involved in neural repair mechanisms and modulates glial cell activity and pain sensitivity [73]. For example, GFAP+ glia in the dorsal root ganglia modulate nociceptive neuronal activity and can release pro-inflammatory cytokines. Peripheral neuropathy is often associated with metabolic disorders such as diabetes. Given MTNR1B’s role in glucose regulation and its association with type 2 diabetes [74], we hypothesize that MTNR1B could indirectly influence the development of peripheral neuropathy through metabolic pathways within glucose/insulin pathways. Another receptor that we found in the context of diabetic polyneuropathy was CCKBR which upon activation can influence the expression of neurotrophic factors that can protect against nerve damage. Finally, we found rare variants in ADORA2B, ADRA1B and HTR6 associated with musculoskeletal pain, especially low back pain.

**Table 6:** Top ranking associations in known pain relevant ion channels & GPCRs.

| Phenotype | code | Gene Symbol | MAF | C<br>H<br>R<br>O<br>M | GENPOS | ID | R<br>E<br>F | A<br>L<br>T | P | OR | case<br>_HO<br>M_<br>REF<br>_CT | case<br>_HE<br>T_<br>R_<br>EF_<br>ALT<br>_CT<br>S | case<br>_T<br>WO<br>_AL<br>T_<br>G<br>ENO<br>_CT<br>S | control_H<br>OM_REF_<br>CT | con<br>trol<br>_H<br>ET_<br>REF_<br>AL<br>T_<br>C<br>TS | con<br>trol<br>_T<br>WO<br>_AL<br>T_<br>GE<br>NO<br>_CT<br>S |
| --- | --- | --- | --- | --- | --- | --- | --- | --- | --- | --- | --- | --- | --- | --- | --- | --- |
| G60.0<br>Hereditary<br>motor and<br>sensory<br>neuropathy | icd.indivi<br>dual.G6<br>00 | CELSR2 | 4.36E-05 | 1 | 109264521 | rs747177120 | A | G | 6.1179E-11 | 156.331 | 163 | 2 | 0 | 140070 | 12 | 0 |
| M54.58 Low<br>back pain<br>(Sacral and<br>sacroccocygeal<br>region) | icd.indivi<br>dual.M5<br>458 | ADORA2B | 4.36E-05 | 17 | 15945264 | rs749875147 | C | A | 2.6278E-10 | 54.234 | 445 | 3 | 0 | 140065 | 17 | 0 |
| G60.0<br>Hereditary<br>motor and<br>sensory<br>neuropathy | icd.indivi<br>dual.G6<br>00 | NTSR2 | 5.00E-05 | 2 | 11660086 | rs149468069 | G | A | 4.6486E-10 | 117.505 | 163 | 2 | 0 | 140068 | 14 | 0 |
| G60.0<br>Hereditary<br>motor and<br>sensory<br>neuropathy | icd.indivi<br>dual.G6<br>00 | ADGRA3 | 8.19E-05 | 4 | 22387944 | rs765533879 | A | G | 2.0373E-09 | 92.5608 | 163 | 2 | 0 | 140064 | 18 | 0 |
| G60.0 Hereditary motor and sensory neuropathy | icd.individual.G600 | MTNR1B | 1.01E-04 | 11 | 92981583 | rs201290817 | G | A | 6.2982E-09 | 76.6157 | 163 | 2 | 0 | 140059 | 23 | 0 |
| G60.0 Hereditary motor and sensory neuropathy | icd.individual.G600 | CELSR3 | 5.96E-05 | 3 | 48651925 | rs139837673 | C | T | 6.933E-09 | 77.1941 | 163 | 2 | 0 | 140062 | 20 | 0 |
| G63.2 Diabetic polyneuropathy | icd.individual.G632 | CCKBR | 6.07E-05 | 11 | 6260043 | rs539605126 | T | G | 1.2426E-08 | 42.5925 | 1104 | 3 | 0 | 140070 | 12 | 0 |
| M54.50 Low back pain (Multiple sites in spine) | icd.individual.M5450 | ADRA1B | 2.77E-05 | 5 | 159972314 | rs911750399 | C | G | 1.8217E-08 | 170.92 | 225 | 2 | 0 | 140077 | 5 | 0 |
| M54.55 Low back pain (Thoracolumbar region) | icd.individual.M5455 | HTR6 | 2.06E-04 | 1 | 19678658 | rs201083688 | C | T | 2.1972E-08 | 59.5318 | 112 | 2 | 0 | 140036 | 46 | 0 |
| G60.0 Hereditary motor and sensory neuropathy | icd.individual.G600 | KCNB2 | 2.767E-05 | 8 | 72937525 | rs199530102 | G | A | 8.9924E-11 | 165.372 | 163 | 2 | 0 | 140070 | 10 | 0 |
| idiopathic neuropathy [ICD group] | icd.group.idiopathic.neuropathy | KCNB2 | 2.767E-05 | 8 | 72937525 | rs199530102 | G | A | 9.7755E-09 | 91.2266 | 298 | 2 | 0 | 140070 | 10 | 0 |
| M06.00 Seronegative rheumatoid arthritis (Multiple sites) | icd.individual.M0600 | KCNE1 | 7.3441E-05 | 21 | 34449526 | rs202036483 | T | C | 3.8337E-09 | 11.6791 | 307 | 0 | 2 | 140048 | 3 | 6 |
| R52.0 Acute pain | icd.individual.R520 | KCNE3 | 2.1284E-05 | 11 | 74457285 | rs199972628 | A | T | 8.5604E-11 | 207.096 | 215 | 2 | 0 | 140073 | 6 | 0 |
| back disorders<br>other [ICD<br>group] | icd.grou<br>p.back.di<br>sorders.<br>other | KCNG2 | 9.5779E-05 | 18 | 79863984 | rs764813108 | C | A | 3.5929E-08 | 61.01 | 210 | 2 | 0 | 140057 | 23 | 0 |
| peripheral<br>neuropathy<br>[Interview<br>UKB:20002] | interview<br>.1255 | KCNG4 | 7.6623E-05 | 16 | 84236998 | rs769378154 | C | T | 2.6477E-08 | 35.4241 | 737 | 3 | 0 | 140066 | 16 | 0 |
| G60.8 Other<br>hereditary and<br>idiopathic<br>neuropathies | icd.indivi<br>dual.G6<br>08 | KCNH3 | 8.5146E-06 | 12 | 49556371 | rs105476937<br>9 | G | A | 1.9246E-08 | 1197.37 | 97 | 1 | 0 | 140069 | 2 | 0 |
| M50.3 Other<br>cervical disk<br>degeneration | icd.indivi<br>dual.M5<br>03 | KCNJ12 | 0.000161<br>82 | 17 | 21415481 | rs201403828 | G | A | 4.6605E-09 | 25.9237 | 958 | 4 | 0 | 139986 | 41 | 0 |
| M54.50 Low<br>back pain<br>(Multiple sites in<br>spine) | icd.indivi<br>dual.M5<br>450 | KCNJ16 | 1.1706E-05 | 17 | 70132016 | 17:70132016<br>:C:G | C | G | 2.7855E-10 | 617.507 | 225 | 2 | 0 | 140079 | 2 | 0 |
| G58.8 Other<br>specified<br>mononeuropathi<br>es | icd.indivi<br>dual.G5<br>88 | KCNJ2 | 5.5339E-05 | 17 | 70176244 | rs759070406 | C | T | 4.0732E-08 | 66.3058 | 302 | 2 | 0 | 140069 | 13 | 0 |
| R52.9 Pain,<br>unspecified | icd.indivi<br>dual.R52<br>9 | SCNN1A | 2.7669E-05 | 12 | 6348126 | rs545954539 | C | T | 4.1384E-08 | 82.3945 | 497 | 2 | 0 | 140074 | 7 | 0 |
| M19.09 Primary<br>arthrosis of<br>other joints (Site<br>unspecified) | icd.indivi<br>dual.M1<br>909 | SCNN1B | 2.8734E-05 | 16 | 23378749 | rs770323212 | C | G | 7.7017E-13 | 721.708 | 130 | 2 | 0 | 140078 | 4 | 0 |
| M13.09<br>Polyarthritis,<br>unspecified (Site<br>unspecified) | icd.indivi<br>dual.M1<br>309 | SCNN1D | 1.2771E-05 | 1 | 1287124 | rs376559305 | G | A | 2.1761E-08 | 696.368 | 92 | 1 | 0 | 140078 | 3 | 0 |

Another important target class for chronic pain are ion channels that play a crucial role in neuronal signal transduction processes. They are involved in the initiation and transmission of pain signals and can influence the excitability of neurons and the release of neurotransmitters [75]. While many ion channels have been implicated in pain sensation for instance, rare variants in Nav1.7 are associated with conditions like erythromelalgia and congenital insensitivity to pain [76], there are few associations within genome wide association studies. A rather unexplored class that we found are the epithelial sodium channels (ENaCs), heterotrimeric ion channels widely distributed throughout various organs, blood vessels and have been implicated in mechano-sensation[77]. Additionally, we found novel variants in these known pain genes that mandate further site-directed mutagenesis and electro-physiological characterization. These include for instance rare non-synonymous, 5’ and 3’ UTR and stop gain variants in potassium channels. An example is a non-synonymous variant rs202036483 in KCNE1 which had two homozygous carriers with multi-site seronegative arthritis. Recent research has highlighted the role of KCNG4 in modulating pain perception, and we found associated variants that expand these observations to other subtypes of KCNG channels.

Most of these rare variant associations are low frequency variants, with a low number of minor allele carriers which will require further confirmation as more cohorts of whole exome sequencing become available.

## Discussion

Here we present a comprehensive genetic analysis of chronic pain within the UK Biobank, we identify many signals within specific chronic pain conditions as well as shared risk factors for chronic pain across conditions.

By using an optimised control cohort, we identify many genome-wide significant hits the chronic pain groups which were previously not reported in broadly applied whole exome sequencing studies in the UK Biobank [31]. We suggest these associations are observed due to minimizing instances of chronic pain within the control group. This hypothesis is further supported by the high level of genetic correlation between the chronic pain conditions, which would mask associations relevant for chronic pain. For instance, we show that in knee pain the previous association within the GDF5 locus is genome-wide significant even when different controls groups are used. Therefore, we recommend that future genetic studies on pain should exclude individuals with other chronic pain conditions from the control group whenever possible. Although this approach may reduce the sample size, the benefit of achieving more precise and meaningful comparisons outweighs the drawback of a smaller sample. In the following paragraphs we have selected results on chronic pain conditions that are still not well understood and involve complex interactions within the central nervous system, with neuroinflammation, central sensitization, and altered neural processing playing significant roles in their pathophysiology. According to our experience, based on the results from this study we found that the definition of the control group is as important as the definition of the cases.

### Shared associations

We looked across a broad range of chronic pain conditions and therefore can find shared genetic risks across the conditions. PLEKHG4B, PRKRA and Mucins were frequently associated with many chronic pain conditions (see supplementary table 3). The interplay between mitochondria, epithelial integrity, Schwann cells, and inflammation involves complex and interconnected biological mechanisms as potential common factors of chronic pain [78] [79]. Linked key pathways such as ROS production, NF-κB signalling, cytokine production, and mitochondrial bioenergetics play central roles in mediating these interactions.

### Epithelial integrity (PLEKHG4B)

PLEKHG4B was associated with 3 non-synonymous variants rs12516846 in fibromyalgia and carpal tunnel syndrome, rs12523402 in Nerve destruction and joint pain, and rs13436090 in rheumatoid arthritis. All the associations showed a protective effect against the pain conditions with a reduced risk with the minor allele. PLEKHG4B is a Rho-guanine nucleotide exchange factor involved in formation of epithelial cell junctions [80] which is highly expressed in the brain, in particular the choroid plexus [45]. The choroid plexus epithelium makes the blood-cerebrospinal fluid barrier, which has been shown to be dysfunctional in many neurological disorders [81]. Other similar rho-guanine nucleotide exchange factors have also been implicated in activity dependent modulation of spinal long term potentiation [82], which may provide an alternative mechanism of PLKHG4B in development of chronic pain. These associations support investigation role of PLEKHG4B in chronic pain.

### Viral response & inflammation (MUC3A, PRKRA, HLA)

Several genes have associated with viral response and inflammation across pain conditions. Among these were Mucins, PRKRA, and the major histocompatibility complex (MHC) region of the genome. Variants within mucins, in particular MUC3A were found across many of the musculoskeletal and soft tissue chronic pain conditions including back pain, pelvic pain, carpal tunnel syndrome and rheumatoid arthritis. Mucins are secreted glycoproteins from epithelial cells which function to protect the epithelial membranes [83]. MUC3A is a membrane bound mucins which can be cleaved to be secreted [84]. MUC3A is primarily secreted in the intestine [85], but has also been found in the knee synovial tissue from epithelial and macrophages [86]. Non-synonymous variant associations within the secreted region of MUC3A: rs73714229 (S172R) and rs73714238 (I2573T) were associated with chronic musculoskeletal and an additional stop gain variant, rs79874934 was associated with back pain. Volin et al. also showed an increased expression of MUC3A in both rheumatoid arthritis and osteoarthritis [86]. We hypothesize that these MUC3A associations influence joint stability and inflammation in these conditions that then contribute to the development of chronic pain. Other whole exome sequencing studies have found associations with ulcerative colitis and MUC3A within different variants in this region (C1019T) [87].

Additionally, we found the mucins MUC16 and MUC4 also associated with multiple pain conditions. Interestingly, these have been shown to undergo cleavage after the activation of a sodium ion channel TRPV4 [88] which is activated in response to noxious stimuli[89]. What the underlying function of mucin release after nociceptor activation is a potential unexplored mechanism of chronic pain.

PRKRA associated with musculoskeletal pain, postherpetic neuralgia and trigeminal neuralgia via rs75862065, and with rheumatoid arthritis via rs77419724 and rs9406386. PRKRA is protein kinase involved in response to dsRNA and potential of triggering apoptosis. It is also a known pain gene from the human phenotype ontologies for limb pain [90]. Inflammation and apoptosis are mechanisms by which PRKRA may lead to chronic pain, particularly in post-viral pain such as post-herpetic neuralgia.

The major histocompatibility complex region (MHC) on chromosome 6 had many associations primarily within seropositive rheumatoid arthritis [31]. The MHC region is one of the most important regions of the genome for immune response including antigen presentation, inflammation and the complement pathway. Interpretation of specific associations in the MHC region is complex due to high levels of linkage disequilibrium, polymorphisms and structural variation [91]. Other studies have shown associations in HLA haplotype differentially between seropositive and seronegative rheumatoid arthritis patients [92].

### Protective rare variants

Protective rare variants can serve as signposts towards proteins and pathways that may be targeted for novel analgesic therapies[93]. We define these as genetic mutations with an odds ratio below 0.7 that have higher frequency of the minor allele in the controls than in the cases. This is a further reason why the definition of control groups plays an important role in the ability to detect such protective rare variants. We focused in our selection on missense and loss of function mutations that generate a testable hypothesis. Since most of these mutations result in a reduced or lost protein activity, studying them can provide insights into the molecular mechanisms. This can be done by experimentally silencing a gene or inhibiting the protein as well as site-directed mutagenesis to replicate the effects of the missense and loss of function mutations. This approach leverages the natural genetic resistance occasionally observed in individuals as known from insensitivity to pain mutations. For peripheral neuropathy we identified a missense mutation in Agrin (Glu728Val) that is known for its role in the neuromuscular junction and inhibition of GABA neurons. It plays a key role in the clustering of acetylcholine receptors (AChRs) at the postsynaptic membrane, which is essential for effective neuromuscular transmission and deletion of agrin from adult motor neurons results in the loss of AChRs that leads to atrophy and withdrawal of nerve terminals from muscle fibers. Also, Autoantibodies against agrin have been identified in patients with myasthenia gravis that extends beyond the neuromuscular junctions to peripheral nerve repair. Further, we found several protective mutations in autoimmune related indications like a LoF mutation in NOTCH4 that can prevent the activation of microglia and the release of pro-inflammatory factors [94]. FKBPL, a negative regulator of angiogenesis that may restore endothelial function and vascular integrity[95]. Since inflammation is a key factor in the development and maintenance of chronic pain via formation of autoantibodies, we found a LoF variant in BTNL2 that regulates T cell responses and upon deletion can disrupt normal immune regulation and contribute to autoimmune disease pathogenesis. Tenascin-XB is an extracellular matrix component that influences cellular processes such as adhesion, migration, and proliferation[96]. Tenascin deficiency is also associated with Ehlers-Danlos syndrome and promoting the activation of TGF-β which is a common feature in many autoimmune diseases [97]. Finally, CHL1 (Close Homolog of L1) was found to be protective for joint pain. Its function has a role in guiding regeneration of motor axons, regulating synaptic coverage of motor neurons and peri somatic innervation hence changing the recovery process after nerve injury [98]. Our major findings in protective rare variants are summarized in the Table 7.

**Table 7:**
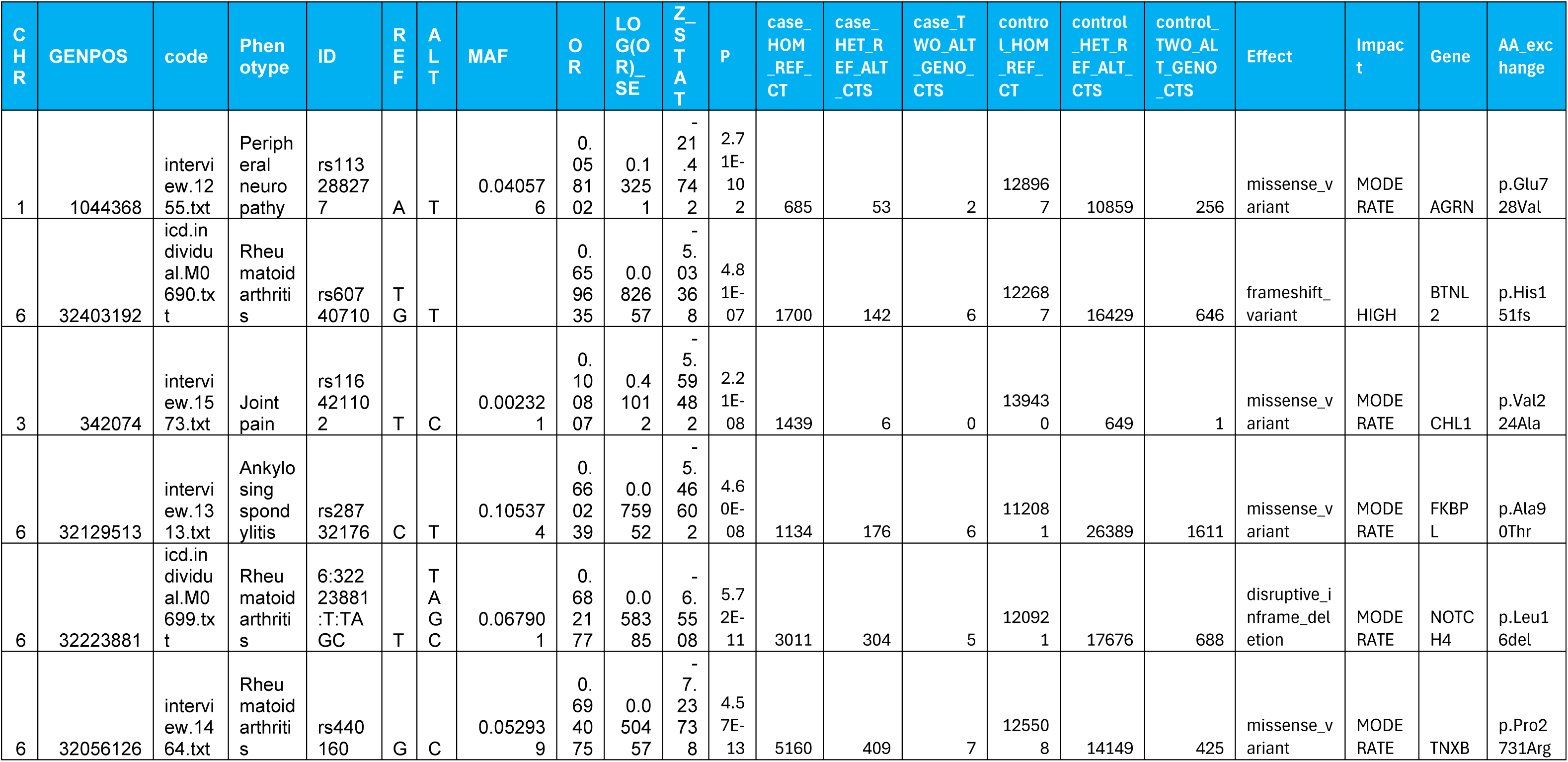
Protective rare variants.

### Strengths and limitations

We have identified rare variants in coding and non-coding regions of the human genome and subsequently used annotations to assess the effect of mutations that are based on models to predict a consequence of a nucleotide change. This works very well for missense and loss of function mutations. However, the functional consequence of rare missense mutations is often unclear especially for novel, unreported variants. Therefore, there is a clear need for experimental validation like site-directed mutagenesis followed by biological comparison of wild type and mutant protein with respect to reported pain scores and their link to pain perception. Since we identified rare variants that usually only have a few carriers in cases and controls the relevance within a broad disease population might be limited. We found high odd ratios for such variants that can be representative to define the pathophysiology and clinical presentation. We think that therapeutic intervention for pathogenic mechanisms found in such genetically defined small pain populations can translate and be efficacious for the broader pain population of the same indication. Common variants from genetic associations usually found in GWAS have low odd ratios and therefore only minor contributions for a specific variant. Aggregation methods like burden tests can also obscure potential therapeutic targets via loss of individual signals, the heterogeneity of diverging effects by missense variants and limited distribution statistics for the saddle point approximation.

Further, the definitions of pain phenotypes are based on self-reported surveys without the presence of a physician or ICD codes are assign by an algorithm based on procedure codes and diagnosis from hospital inpatient records. While we find associations in our work, the specificity is inevitably not comparable to a physician-based diagnosis.

The research did not specifically consider self-reported ethnicity of participants, but this was indirectly addressed by incorporating genetic principal components as covariates in the analysis. Since the UK Biobank is mostly white British the effect of including these participants is likely small but may have a larger impact on rarer variants with differing minor allele frequency between ethnic groups. This may lead to an increase in false positive associations, and therefore our interpretation of results has considered the allele frequencies within the respective populations.

## Data Availability

This research has been conducted using data from UK Biobank, a major biomedical database under project ID 64765.

## Summary

This study is a comprehensive analysis of the genetic background especially rare variants of chronic pain phenotypes within the whole UK Biobank including array and whole exome sequencing data. By using optimized control cohorts, we can consider shared genetic contributors to chronic pain across many conditions and investigate pleiotropic effects. We leveraged rare variant analysis and identified a realm of novel genome-wide significant associations with p-values below 5E-08 of rare protective and causal variants for all pain conditions defined by ICD-10 codes and novel pain questionnaire data within the UK Biobank. Our findings implicate novel genes and regulatory pathways for enigmatic pain conditions, paving the way for future research on personalized pain management and therapeutic targets. We presented several selected examples of our findings for complex regional pain syndrome and Fibromyalgia suggesting novel mechanistic insights into their autoimmune and blood vessel pathophysiology. Cross analysis over more than 300 pain conditions revealed autoimmune related mechanisms as the most common driver in chronic pain conditions. We also identified novel protective rare variants that may offer insights into potential roles in pain resilience mechanisms, pain sensitivity and modulation. Finally, epistasis analysis revealed significant interactions between rare variants in different genes, highlighting their cooperative effects on the penetrance of pain modulatory mechanisms.

## Acknowledgements

UK Biobank is a large-scale biomedical database and research resource containing genetic, lifestyle and health information from half a million UK participants. This research has been conducted using data from UK Biobank, a major biomedical database under project ID 64765. The UK Biobank database received ethical approval from the North-West Haydock Research Ethics Committee (REC reference 21/NW/0157) and participants gave informed consent. We would like to thank all participants of the UK Biobank cohort who have provided necessary genetic and phenotypic information.

## Supplementary Material

Figure S1: Pain by region over last 3 months. Each point shows the median for a specific body area, with max taking the maximum value across all areas where the value was not “no pain”.

Supplementary Table 1: Phenotypic descriptions within the chronic pain subset.

Supplementary Table 2: Pain relevant genesets from MSIGdb

Supplementary Table 3: All genome wide significant results across all cohorts.

Supplementary Table 4: Phenotypic descriptions of all cohorts.

Supplementary Table 5: HPO terms

---

Fine-mapping Chr 11 (SIAE):

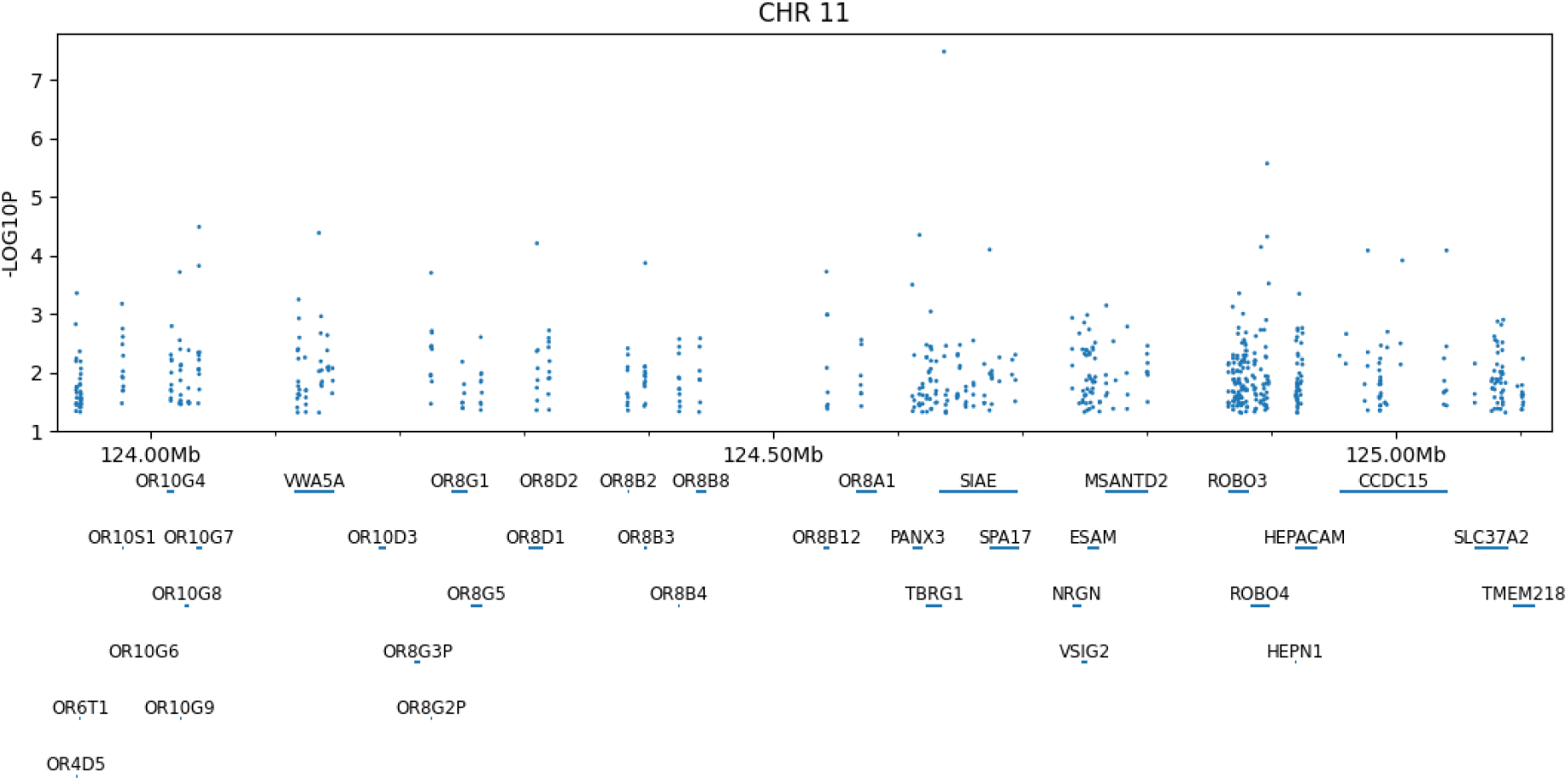

